# Trends in weight gain recorded in English primary care before and during the Coronavirus-19 pandemic: an observational cohort study using the OpenSAFELY platform

**DOI:** 10.1101/2023.04.01.23287538

**Authors:** Miriam Samuel, Robin Y Park, Sophie V Eastwood, Fabiola Eto, Caroline E Morton, Daniel Stow, Sebastian Bacon, Amir Mehrkar, Jessica Morley, Iain Dillingham, Peter Inglesby, William J Hulme, Kamlesh Khunti, Rohini Mathur, Jonathan Valabhji, Brian MacKenna, Sarah Finer, The OpenSAFELY Collaborative

## Abstract

**Background:** We investigated which clinical and sociodemographic characteristics were associated with unhealthy patterns of weight gain amongst adults living in England during the pandemic.

**Methods:** With the approval of NHS England we conducted an observational cohort study of Body Mass Index (BMI) changes between March 2015 and March 2022 using the OpenSAFELY-TPP platform. We estimated individual rates of weight gain before and during the pandemic, and identified individuals with rapid weight gain (>0·5kg/m^2^/year) in each period. We also estimated the change in rate of weight gain between the prepandemic and pandemic period and defined extreme-accelerators as the ten percent of individuals with the greatest increase (>1·84kg/m^2^/year). We estimated associations with these outcomes using multivariate logistic regression.

**Findings:** We extracted data on 17,742,365 adults (50·1% female, 76·1% White British). Median BMI increased from 27·8kg/m^2^ [IQR:24·3-32·1] in 2019 (March 2019 to February 2020) to 28·0kg/m^2^ [24·4-32·6] in 2021. Rapid pandemic weight gain (n=3,214,155) was associated with female sex (male vs female: aOR 0·76 [95%CI:0·76-0·76]); younger age (50-59-years vs 18–29-years: aOR 0·60 [0·60-0·61]); White British ethnicity (Black Caribbean vs White British: aOR 0·91 [0·89-0·94]); deprivation (least-deprived-IMD-quintile vs most-deprived: aOR 0·77 [0·77-0·78]); and long-term conditions, of which mental health conditions had the greatest effect (e.g. depression (aOR 1·18[1·17-1·18])). Similar characteristics increased risk of extreme acceleration (n=2,768,695).

**Interpretation:** We found female sex, younger age, deprivation and mental health conditions increased risk of unhealthy patterns of pandemic weight gain. This highlights the need to incorporate sociodemographic, physical, and mental health characteristics when formulating post-pandemic research, policies, and interventions targeting BMI.

**Funding:** NIHR

## Background

Obesity and rapid weight gain^1^ are established risk factors for non-communicable diseases and have emerged as independent risk factors for severe disease following Coronavirus-19 (COVID-19) infection.^2–5^ In March 2020, restrictions imposed to reduce COVID-19 transmission resulted in profound societal changes that impacted many health behaviours, including physical activity and nutrition.^3,6–8^ However, the suspension of population health surveys, such as the Health Survey for England, during the pandemic^9^ resulted in a lack of quantitative data on weight changes.

A systematic review of observational studies reported a modest increase in adult weight during the pandemic, but analyses were limited by small sample size and non-representative samples.^10^ Studies using routinely collected healthcare records have been limited to populations without universal access to healthcare,^11–13^ or individuals with long-term conditions (LTCs).^14^ In these settings, women,^11,12^ young adults,^11,12^ those living in deprivation,^13^ and those with LTCs,^13,14^ including depression,^13^ were at greatest risk of weight gain during the pandemic.^7^ However, these findings have not been replicated in a population-representative cohort. Additionally, groups such as young adults, were at increased risk of weight gain even prior to the pandemic.^15^ Therefore, a comparison of individual rates of weight gain, before and after the onset of the pandemic is required to understand the specific impact of the pandemic and identify individuals who had the greatest acceleration in their rate of weight gain during the pandemic (extreme-accelerators). Body Mass Index (BMI) data, a measure of weight adjusted for height, recorded in English primary care Electronic Health Records (EHRs) were used to estimate rates of weight gain (BMI trajectories) before the pandemic,^15^ and provide a source of nationally representative data available during the pandemic.^16^

Using routinely collected English primary care records we aimed to: (i) describe population-level changes in BMI during the pandemic and (ii) estimate individual-level rates of weight gain before and during the pandemic to identify how (iia) risk of rapid weight gain (>0·5kg/m^2^/year) in the pre-pandemic and pandemic periods and (iib) risk of extreme acceleration in the rate of weight gain between the prepandemic and pandemic period varied by sociodemographic and clinical characteristics. The results of these analyses will inform targeted policy to deliver weight loss interventions in the post-pandemic recovery.

## Methods

### Data Source

All data were linked, stored and analysed securely within the OpenSAFELY-TPP platform, https://opensafely.org/, containing pseudonymised data on approximately 40% of the English population, including coded diagnoses, medications and physiological parameters. No free text data are included. Detailed pseudonymised patient data is potentially re-identifiable and therefore not shared. The study was approved by the London School of Hygiene & Tropical Medicine Ethics Board (reference 26536). An information governance statement is included (Appendix 3).

### Study population

We extracted data on all male and female adults aged ≥18 to <90 years who had been registered with a primary care practice using TPP EHR software for at least one year prior to 1st March 2022.

### Study Outcomes

We used changes in Body Mass Index (BMI, a measure of weight adjusted for height), as a proxy for weight change. BMI is recorded in primary care records during routine health checks, disease monitoring, and opportunistically. We extracted BMI data using recorded weight (in kilograms) and height (in metres), or directly from recorded BMI values (in kg/m^2^) between 1st March 2015 and 1st March 2022, from the point an individual reached 18 years of age. Individuals who joined the OpenSAFELY-TPP database after 1st March 2015 had BMI data extracted between their date of first registration with the GP practice and 1st March 2022. Recordings taken within healthcare settings and self-reported values are included in EHRs and cannot be differentiated using clinical codes. We extracted monthly values per individual and took the most recent in instances where multiple values per calendar month were present. Extreme values (BMI<15kg/m^2^ and BMI>65kg/m^2^) were omitted from all quantitative analyses related to BMI values to censor erroneous results and exclude extremely overweight and underweight individuals.

### Population level trends in BMI recording activity and median BMI

We examined the proportion and characteristics of the population having at least one BMI value recorded in the year beginning March 2019 (2019), March 2020 (2020), and March 2021(2021) to characterise how the pandemic had influenced BMI data availability. We then calculated the population-level median and interquartile range (IQR) of the recorded BMI values for each of the years above, using the median per year, where multiple values were available for a single individual.

### Individual level BMI trajectories in pre-pandemic and pandemic periods

We first estimated individual level BMI trajectories as rates of BMI change per year (δ) in kg/m^2^/year, before the onset of COVID-19 pandemic (δ-prepandemic) and after the onset of the pandemic (δ-pandemic). To calculate δ-prepandemic we randomly selected one pair of BMI measurements from individuals who had a BMI value recorded in each of the following time windows (period-1: March 2015 - February 2018; and period-2: March 2018 - February 2020), and calculated the rate of BMI change/year assuming a linear trend following established methods.^15^ To calculate δ-pandemic we replicated this method, with a random pair of BMI measures from each of the following time windows (period-2: March 2018 - February 2020; and period-3: March 2020 - February 2022). (Appendix 1) Individuals with known cancer and those underweight (BMI <18·5kg/m^2^) in the prepandemic period were excluded from these analyses as their patterns of weight change were likely to differ from the general population. The most extreme 0·05% of values of δ-prepandemic and δ-pandemic (a positive or negative change of >6kg/m^2^/year) were censored to reduce the impact of erroneous results. Individuals gaining >0·5kg/m^2^/year were classified as experiencing rapid weight gain.^1^ We estimated associations between socio-demographic and clinical characteristics and rapid weight gain before and during the pandemic using descriptive statistics and calculating odds adjusted for age, sex, ethnicity and IMD.

### Defining a population of extreme accelerators

To identify individuals who had experienced the greatest acceleration in their rate of weight gain between the prepandemic and pandemic period, we first estimated individual-level changes in rate of weight gain (δ-change) as the difference in the prepandemic and pandemic rates of weight gain (δ-change = δ-pandemic - δ-prepandemic) (see Appendix 1 for a more detailed description of the methods). A positive δ-change indicated the rate of weight gain increased or, if patients were losing weight prepandemic, the rate of weight loss slowed. We then defined the ten percent of the study population experiencing the greatest acceleration in their rate of weight gain (δ-change >= 1·84kg/m^2^/year) as ‘extreme accelerators’. We estimated associations between socio-demographic and clinical characteristics and extreme acceleration using descriptive statistics and calculating odds adjusted for age, sex, ethnicity and IMD.

### Covariates

Covariates were chosen by clinical consensus, guided by data availability and known potential predictors of BMI change. Covariates included age (18-29 years, 30-39 years, 40-49 years, 50-59 years, 60-69 years, 70-79 years, 80 ≤ 90 years), sex (female or male); ethnicity, based on the 2001 UK Census definitions (White British, White Irish, Other White, Black African, Black Caribbean, Other Black, Indian, Pakistani, Bangladeshi, Chinese, Other Asian, Mixed White/Black African, Mixed White/Black Caribbean, Mixed White/Asian, Other); deprivation using the most recent patient postcode-derived Index of Multiple Deprivation (IMD; by quintiles from those living in the most deprived 20% of households to the least deprived 20%); and the presence or absence of the following LTCs ever-recorded: hypertension; type 1 diabetes (T1D), type 2 diabetes (T2D), asthma, chronic obstructive pulmonary disease (COPD), anxiety and depression, serious mental illness (SMI: psychosis or bipolar disorder), learning difficulties, dementia, cardiovascular disease (CVD: chronic cardiac conditions), and stroke and transient ischaemic attack (Stroke and TIA).

### Statistical Models

We used complete case analysis (inclusion of individuals with all baseline covariate data) in all statistical models. This is consistent with previous studies using English primary care data.^17^ As this is a descriptive study, multiple imputation techniques were not employed as we were not trying to gain an unbiased estimate of a single exposure-outcome association, and therefore did not have an analytic model around which to build an imputation procedure. Additionally, the missing at random assumption (required for imputation) was unlikely to hold, e.g. individual IMD linked to residence is less likely to be recorded for individuals in unstable accommodation. All individuals had complete data for age and sex as these were part of the study inclusion criteria, and clinical covariates (such as T2D) which were identified based on the presence/absence of specified codes in the data. Therefore, ethnicity and IMD were the only variables with missing data.

For each of the estimated individual-level outcomes (rapid weight gain prepandemic, rapid weight gain pandemic and extreme acceleration in rate of weight gain), we compared the baseline characteristics of the population with and without the outcome. We also compared baseline characteristics and outcomes between the complete case sample and the entire population, including those with missing ethnicity and deprivation data, to identify differences in individuals with and without missing data.

We used logistic regression to explore associations between the sociodemographic and clinical covariates and the estimated outcomes. Models were adjusted separately for age, sex, IMD and ethnicity and then in multivariable models adjusted for age and sex; age, sex and IMD; age, sex and ethnicity; and age, sex, IMD and ethnicity.

### Subgroup Analyses

We investigated whether estimated associations between the covariates and extreme acceleration in rate of weight gain persisted in populations stratified by age group (18-39 years, 40-59 years, and 60-79 years), sex, IMD (IMD quintile 1 and IMD quintile 5) and ethnicity (Black and South Asian (Bangladeshi, Indian, Pakistani)). The ethnicities were grouped into broader groups (White, Black, South Asian, Chinese and other, and Mixed ethnicity) for subgroup analyses to reduce risk of disclosure from rare events.

Data management was done with Python 3·8 and SQL, and analysis was done using R 4·0. Prevalence counts are rounded to the nearest five to reduce risk of disclosure. All code for data management and analysis, as well as codelists is shared openly for review and re-use under MIT open license, available at https://github.com/opensafely/BMI-and-Metabolic-Markers.

### Patient and public involvement

OpenSAFELY has a publicly available <u>website</u> through which we invite patients or members of the public to contact us about this study or the broader OpenSAFELY project.

## Results

Data were extracted for 17,742,365 adults meeting study inclusion criteria of whom 50·07% were female and 76·1% were of white British ethnicity (Table 1). Figure 1 demonstrates the population contributing data to each stage of the analysis.

**Table 1.** Baseline characteristics of the study population at each stage of the analysis.

|  |  |  |  |  |  |  |
| --- | --- | --- | --- | --- | --- | --- |
|  | **Total Study Population\*** | **Population trends in median BMI\*\*** |  | **Individuals level rates of BMI\*\* change (δ)** |  |  |
|  |  | **Year beginning March 2019** | **Year beginning March 2021** | **δ-prepandemic** | **δ-pandemic** | **δ-change** |
|  | **N (%)** | **N (%)** | **N (%)** | **N (%)** | **N (%)** | **N (%)** |
| **Total** | 17,742,365 | 4,422,295 | 5,094,590 | 3,966,500 | 3,214,155 | 2,768,695 |
| **Age Group (years)** |  |  |  |  |  |  |
| 18-29 | 3,062,705 (17.3) | 454,245 (10.3) | 580,335 (11.4) | 274,540 (6.9) | 267,605 (8.3) | 161,655 (5.8) |
| 30-39 | 3,138,645 (17.7) | 469,950 (10.6) | 579,610 (11.4) | 466,285 (11.8) | 341,870 (10.6) | 253,415 (9.2) |
| 40-49 | 2,953,105 (16.6) | 591,250 (13.4) | 735,120 (14.4) | 545,760 (13.8) | 415,805 (12.9) | 310,045 (11.2) |
| 50-59 | 3,176,680 (17.9) | 823,905 (18.6) | 958,860 (18.8) | 757,830 (19.1) | 603,670 (18.8) | 479,655 (17.3) |
| 60-69 | 2,495,010 (14.1) | 839,940 (19.0) | 980,770 (19.2) | 771,890 (19.5) | 642,120 (20.0) | 573,030 (20.7) |
| 70-79 | 1,997,575 (11.3) | 836,005 (18.9) | 885,645 (17.4) | 757,405 (19.1) | 635,175 (19.8) | 649,920 (23.5) |
| 80-89 | 918,640 (5.2) | 406,995 (9.2) | 374,250 (7.3) | 392,785 (9.9) | 307,910 (9.6) | 340,975 (12.3) |
| **Sex** |  |  |  |  |  |  |
| Female | 8,883,720 (50.1) | 2,591,225 (58.6) | 2,943,330 (57.8) | 2,381,345 (60.0) | 1,898,510 (59.1) | 1,612,850 (58.2) |
| Male | 8,858,645 (49.9) | 1,831,065 (41.4) | 2,151,260 (42.2) | 1,585,155 (40.0) | 1,315,645 (40.9) | 1,155,845 (41.8) |
| **Index of Multiple Deprivation** |  |  |  |  |  |  |
| 1 (most deprived) | 3,356,720 (19.4) | 868,410 (20.1) | 1,011,785 (20.2) | 836,480 (21.1) | 686,100 (21.3) | 592,805 (21.4) |
| 5 (least deprived) | 3,240,120 (18.7) | 777,720 (18.0) | 905,350 (18.1) | 692,065 (17.4) | 549,495 (17.1) | 472,800 (17.1) |
| Missing | 431,235 (2.4) | 108,220 (2.5) | 89,270 (1.8) | 93,545(2.2) | 79,215(1.9) | 57,435(2.1) |
| **Ethnicity** |  |  |  |  |  |  |
| White British | 12,299,145 (76.1) | 3,446,610 (81.1) | 4,017,955 (81.9) | 3,275,985 (82.6) | 2,631,235 (81.9) | 2,306,865 (83.3) |
| White Irish | 93,035 (0.6) | 24,460 (0.6) | 27,760 (0.6) | 21,500 (0.5) | 17,590 (0.5) | 15,475 (0.6) |
| Other White | 1,540,215 (9.5) | 254,570 (6.0) | 293,165 (6.0) | 211,145 (5.3) | 172,260 (5.4) | 131,960 (4.8) |
| Indian | 501,025 (3.1) | 129,905 (3.1) | 139,620 (2.8) | 115,585 (2.9) | 99,750 (3.1) | 82,245 (3.0) |
| Pakistani | 358,585 (2.2) | 99,095 (2.3) | 111,395 (2.3) | 99,045 (2.5) | 80,400 (2.5) | 68,000 (2.5) |
| Bangladeshi | 83,300 (0.5) | 23,755 (0.6) | 25,350 (0.5) | 21,230 (0.5) | 18,965 (0.6) | 15,025 (0.5) |
| Chinese | 115,825 (0.7) | 12,910 (0.3) | 15,860 (0.3) | 10,185 (0.3) | 8,245 (0.3) | 6,395 (0.2) |
| Other Asian | 264,080 (1.6) | 62,390 (1.5) | 65,660 (1.3) | 51,865 (1.3) | 46,590 (1.5) | 36,420 (1.3) |
| Black Caribbean | 95,140 (0.6) | 29,520 (0.7) | 32,595 (0.7) | 27,170 (0.7) | 23,050 (0.7) | 23,970 (0.9) |
| Black African | 219,880 (1.4) | 46,435 (1.1) | 49,330 (1.0) | 37,745 (0.9) | 32,875 (1.0) | 20,330 (0.7) |
| Other Black | 89,040 (0.6) | 21,265 (0.5) | 22,845 (0.5) | 17,295 (0.4) | 15,020 (0.5) | 11,300 (0.4) |
| Mixed White/Black Caribbean | 49,930 (0.3) | 11,655 (0.3) | 13,065 (0.3) | 10,610 (0.3) | 8,840 (0.3) | 6,960 (0.2) |
| Mixed White/Black African | 39,065 (0.2) | 7,720 (0.2) | 8,375 (0.2) | 6,400 (0.2) | 5,460 (0.2) | 3,935 (0.1) |
| Mixed White/Asian | 42,740 (0.3) | 8,530 (0.2) | 9,240 (0.2) | 7,055 (0.2) | 5,945 (0.2) | 4,445 (0.2) |
| Other Mixed | 82,940 (0.5) | 15,515 (0.4) | 17,055 (0.3) | 12,525 (0.3) | 10,795 (0.3) | 7,950 (0.3) |
| Other | 286,925 (1.8) | 53,955 (1.3) | 57,620 (1.2) | 41,155 (1.0) | 37,130 (1.2) | 27,420 (1.0) |
| Missing | 1,581,510 (8.9) | 174,020 (3.9) | 187,700 (3.7) | 148,055(3.5) | 121,040(2.9) | 7380(3.2) |
| **Long Term Condition** |  |  |  |  |  |  |
| Hypertension | 3,558,405 (20.1) | 2,673,785 (60.5) | 1,837,260 (36.1) | 1,637,165 (41.3) | 1,436,070 (44.7) | 1,399,300 (50.5) |
| Type 2 Diabetes | 1,231,455 (6.9) | 887,870 (20.1) | 867,735 (17.0) | 820,390 (20.7) | 793,770 (24.7) | 805,315 (29.1) |
| Type 1 Diabetes | 93,505 (0.5) | 55,640 (1.3) | 59,420 (1.2) | 55,810 (1.4) | 51,250 (1.6) | 47,910 (1.7) |
| Learning Disability | 105,855 (0.6) | 65,265 (1.5) | 61,110 (1.2) | 54,580 (1.4) | 55,750 (1.7) | 49,865 (1.8) |
| Depression | 3,592,080 (20.2) | 1,140,545 (25.8) | 1,233,900 (24.2) | 1,105,540 (27.9) | 887,240 (27.6) | 777,355 (28.1) |
| Dementia | 134,080 (0.8) | 60,785 (1.4) | 48,460 (0.9) | 58,670 (1.5) | 47,225 (1.5) | 48,415 (1.8) |
| Serious Mental Illness | 195,450 (1.1) | 118,010 (2.7) | 116,055 (2.3) | 100,025 (2.5) | 101,395 (3.2) | 88,375 (3.2) |
| COPD*** | 531,475 (3.0) | 261,870 (5.9) | 308,030 (6.0) | 299,095 (7.5) | 227,935 (7.1) | 245,510 (8.9) |
| Asthma | 2,930,645 (16.5) | 871,240 (19.7) | 1,040,870 (20.4) | 939,925 (23.7) | 709,730 (22.1) | 638,485 (23.1) |
| Cardiovascular Disease | 1,087,320 (6.1) | 578,685 (13.1) | 588,970 (11.6) | 560,030 (14.1) | 487,915 (15.2) | 498,305 (18.0) |
| Stroke and TIA**** | 465,645 (2.6) | 229,915 (5.2) | 226,455 (4.5) | 219,360 (5.5) | 187,580 (5.8) | 190,500 (6.9) |
| Cancer | 905,475 (5.1) | 348,225 (7.9) | 348,585 (6.8) | 0 | 0 | 0 |
| * Data were extracted on all adults aged $\geq 18$ to $\leq 90$ years who had been registered with a TPP practice for at least one year prior to 1st March 2022<br>** Body Mass Index (BMI)<br>*** Chronic Obstructive Pulmonary Disease (COPD)<br>**** Transient Ischaemic Attack (TIA) | | | | | | |

**Figure 1.**
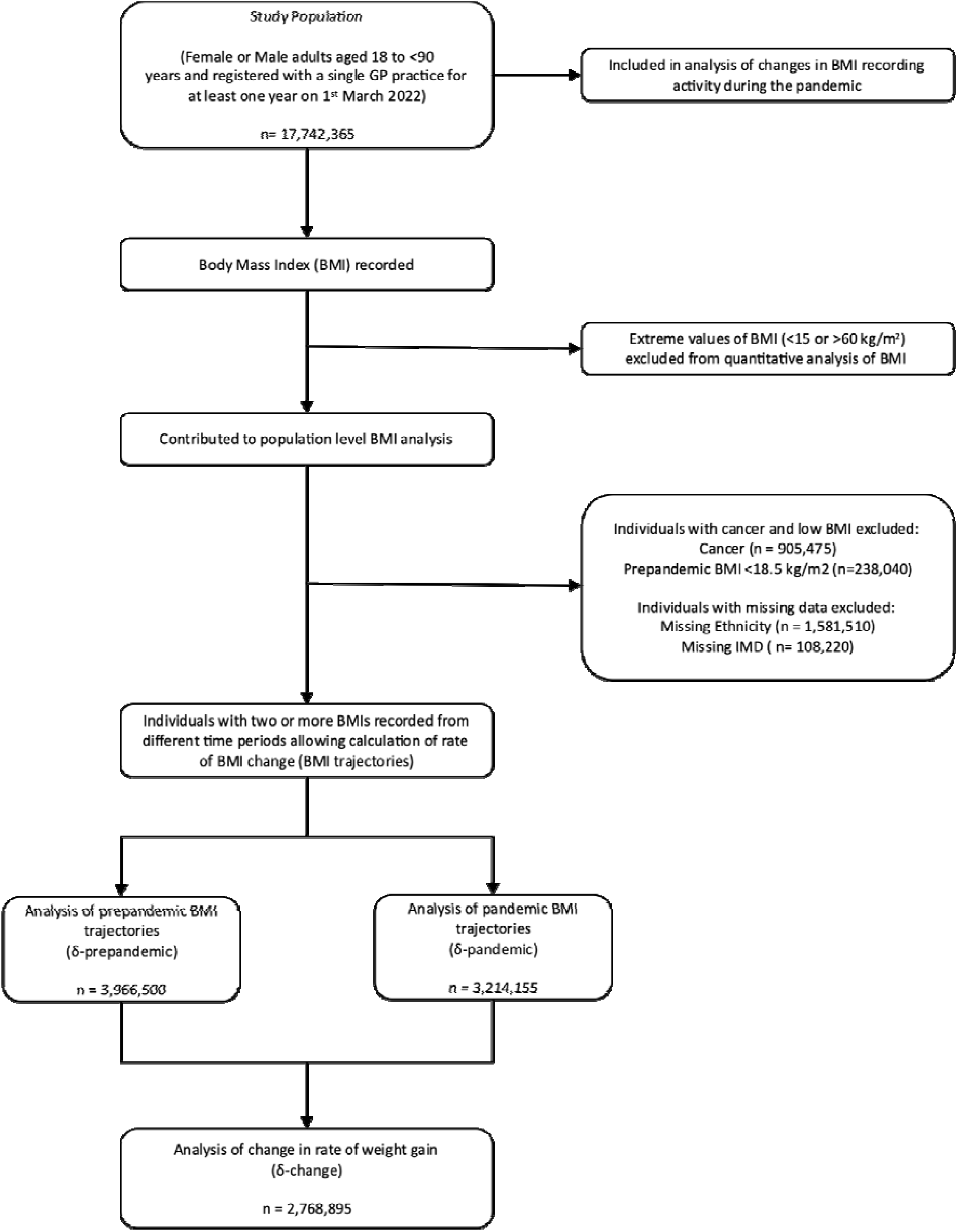
Study Population Flow Chart.

### Population level trends in median BMI

The median recorded BMI increased from 27·8kg/m^2^ [IQR: 24·3-32·1kg/m^2^] in 2019 to 28·0kg/m^2^ [24·4-32·6] in 2021. An increase in median BMI was seen in all subgroups, except the oldest age group (aged 80-89), people with dementia, and people with T2D. (Figure 2)

**Figure 2.**
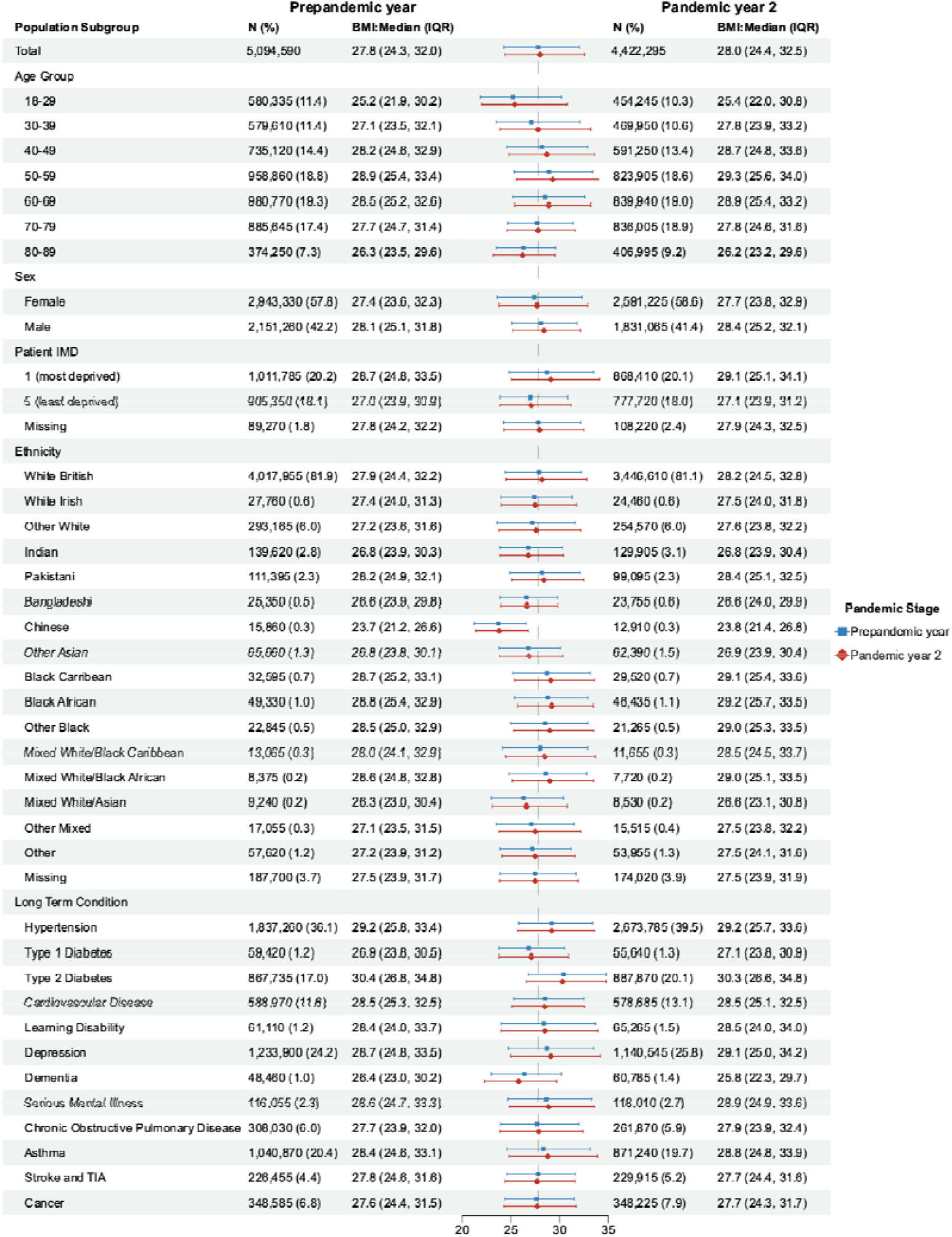
Average Body Mask index (BMI^*^) recorded in the routine healthcare records of adults living in England during the prepandemic year (commencing March 2019) and pandemic year 2 (commencing March 2021). *BMI (in kilograms/metre^2^) were calculated from recorded weight (kg) and height (m) measures, or directly from recorded BMI values

The proportion and characteristics of the population having a BMI recorded changed during the pandemic. BMI was recorded in 31·3% (95% CI: 31·3-31·3) of the total study population in 2019, dropping to 18·7% in 2020 before recovering to 25·0% in 2021, with higher proportions seen amongst individuals with LTCs in all the study periods (e.g. T2D: 82·5% in 2019, 72·2% in 2021). (Supplementary Table 1)

### Individual level BMI trajectory analyses

#### Rapid weight gain (> 0·5kg/m^2^/year) in prepandemic and pandemic time periods

We investigated individual BMI trajectories to identify the groups at greatest risk of rapid weight gain amongst adults with BMI measures recorded in their EHRs. The average rate of weight gain was 0·06kg/m^2^/year (SD:1·20) during the pandemic and 0·08kg/m^2^/year (SD:1·05) prepandemic (Supplementary Table 2). We found 29·2% of individuals gained weight rapidly during the pandemic, compared to 26·9% prepandemic. Figure 3 demonstrates the estimated risk and adjusted odds of rapid weight gain before and after the onset of the pandemic. The risk of rapid weight gain during the pandemic (δ-pandemic) was associated with age, sex, deprivation, ethnicity and a history of LTCs. Adults aged 18-29 years had the highest risk (44·2%), all other age groups had lower risk and adjusted odds (e.g. 50-59-year-olds 30·1%: aOR 0·60 [0·60-0·61]). Male sex reduced the risk and adjusted odds (23·9% male vs 32·9% female: aOR 0·76 [0·76-0·76]), as did living in the least deprived quintile (26·1% IMD5 vs 32·6% IMD1; aOR 0·77 [0·77-0·78]). White British people had the highest adjusted odds of rapid weight gain compared to all other ethnicities; in comparison, Black Caribbean people had the greatest risk (32·4%) but a lower adjusted odds (aOR 0·91[0·89-0·94]). When comparing individuals with and without specific LTCs, T2D was the only LTC associated with a reduction in the estimated risk and adjusted odds (20·73%, aOR 0·71 [0·71-0·72]). Individuals with all the other studied LTCs had an increased estimated risk, with the greatest risk amongst those with learning difficulties (36·3%: aOR 1·10 [1·08-1·12]), SMI (35·2%: aOR 1·23[1·21-1·24]) and depression (33·4%: aOR 1·18 [1·17-1·18]). Risk factors for rapid weight gain before the pandemic were similar (Figure 3).

**Figure 3.**
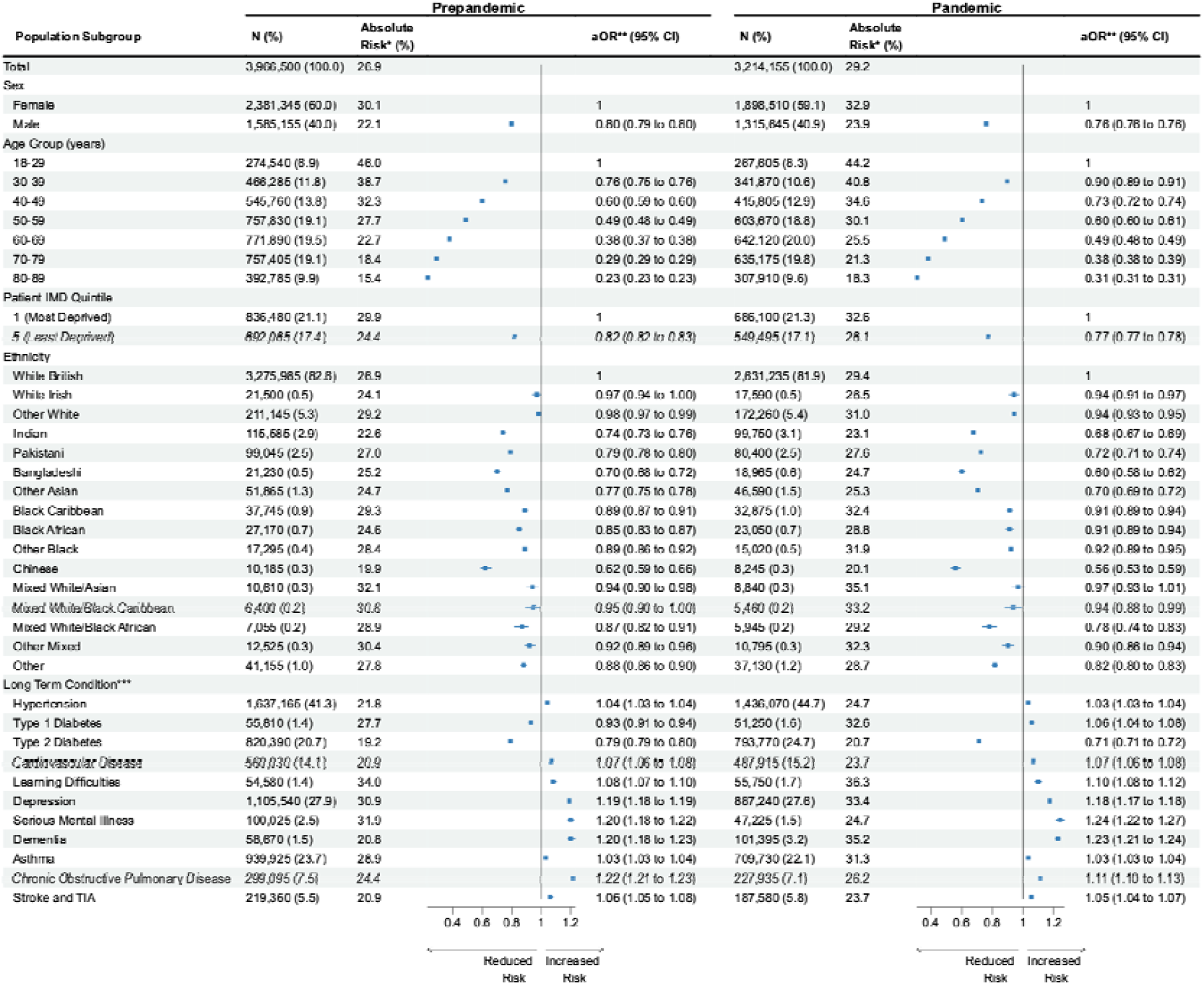
Estimated Risk of Rapid Weight Gain (> 0.5 kg/ m ^2^/year) amongst adults living in England before and after the onset of the COVID-19 pandemic. *Absolute Risk of Rapaid Weight Gain **aOR-Odds Ratio mutually adjusted for sex, age, ethnicity and MD ***aOR for Long Term Conditions presented in comparison to a reference group of Individuals undiagnosed for the respective condition

#### Extreme acceleration in rate of weight gain between the prepandemic and pandemic

To define the population experiencing the greatest increase in their rate of weight gain between the prepandemic and pandemic periods (extreme accelerators), we first estimated the change in rate of weight gain in n=2,768,695 individuals who had prepandemic and pandemic BMI trajectory data (δ-change = δ-pandemic - δ-prepandemic). We found a wide distribution in δ-change (Supplementary Figure 2 in appendix 2), half the population had a slight reduction in their rate of weight gain (median δ-change = -0·02kg/m^2^/year), while 40% of the population experienced an acceleration of more than 0·27kg/m^2^/year. The ten percent of the study population with the greatest acceleration in their rate of weight gain after the onset of the pandemic (δ-change >= 1·84kg/m^2^/year, appendix 2) were classified as extreme accelerators. Consistent with our findings on risk of rapid weight gain, women, younger adults, and individuals living in the most deprived quintiles were at greatest risk of being an extreme accelerator. Living with T1D reduced the adjusted odds (9·4%, aOR 0·87 [0·85-0·89]). All other LTCs increased the adjusted odds, with mental health conditions, including depression, SMI and dementia, having the largest estimated effect. (Figure 4)

**Figure 4.**
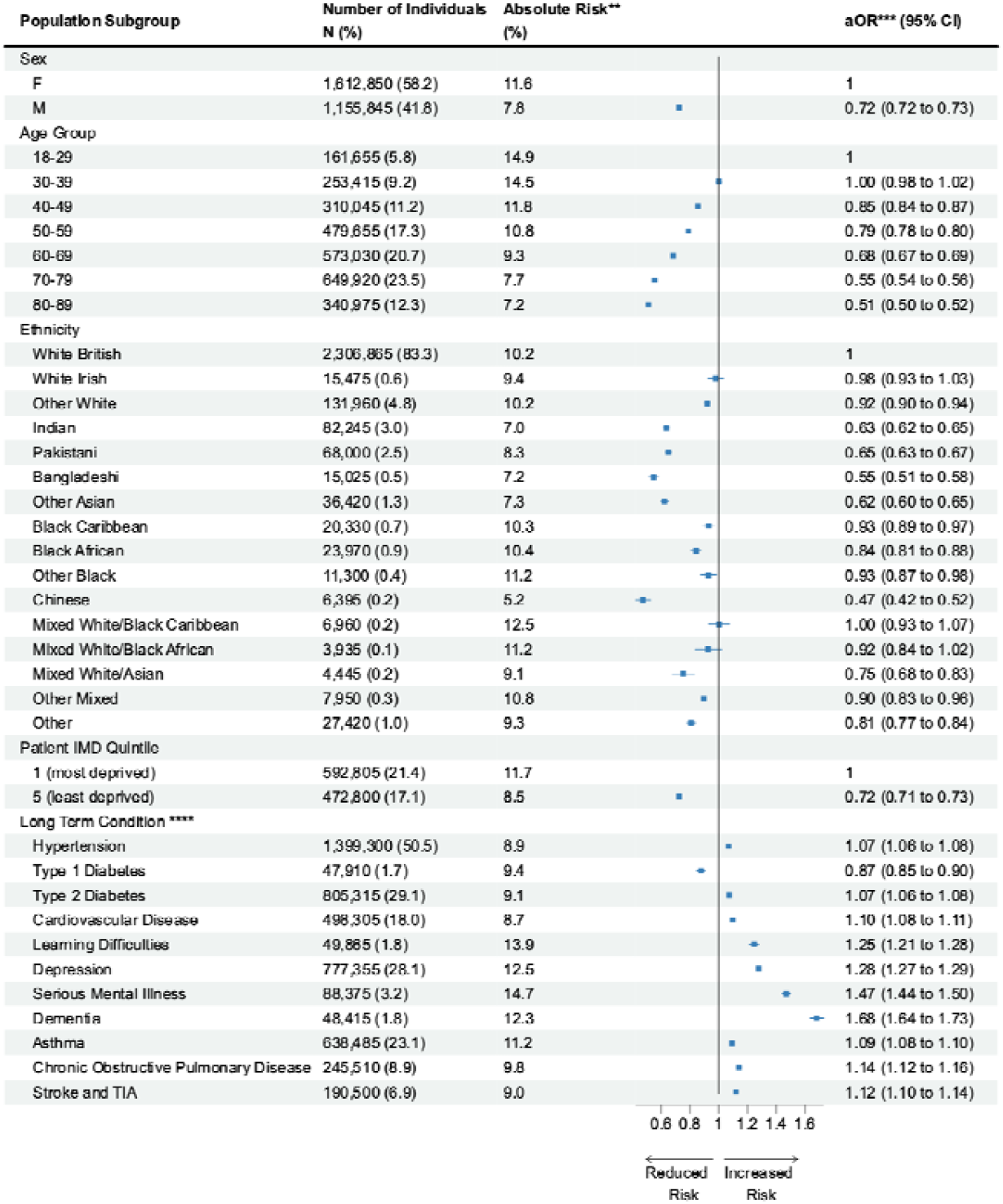
Estimated risk of extreme acceleration^*^ in the rate of weight gain between the prepandemic and pandemic period in 2,786,695 adults living in England. *Extreme accolerators defined as the 10% of the population experiencing the greatost Increase in thelr rate of weight gain between the prepandemic and pandemic period ** Absolute Risk of extreme acceleration in rate of weight gain *** aOR: Odds Ratio mutually adjusted for sex, age, ethnicity and Index of Multiple Deprivation **** aOR for Long-term conditions presented In comparison to a reference group of Individuals undiagnosed for the respective condition

#### Sensitivity and Subgroup Analyses

We compared our findings in the complete case sample to analyses conducted in the entire population, including those with missing ethnicity and deprivation data, and found our results to be consistent in both groups. We found that most of the estimated associations between the risk of extreme acceleration in rate of weight gain and age, sex, IMD and LTCs persisted in the stratified analyses, but there were some exceptions, e.g. amongst Black individuals, deprivation was not associated with risk (IMD5 vs IMD1: aOR 1·04[0·92-1·18]); and amongst those living in the least deprived quintiles, Black individuals were at higher risk than Whites (Black vs White: aOR 1·24 [1·10-1·39]). There was consistent evidence that most LTCs increased the estimated risk of a rapid acceleration in rate of weight gain, with mental health conditions continuing to have the greatest effect.

Associations with T2D were less consistent, a history of T2D continued to have an estimated protective affected amongst South Asian (aOR 0·84 [0·80-0·87]) and Black (aOR 0·88 [0·83-0·94]) individuals, but increased the estimated risk in other subgroups including older adults (aged 60-79: aOR 1·11 [1·09-1·12]) and the least deprived IMD quintile (aOR 1·17 [1·15-1·20]) (Supplementary Table 2-6).

## Conclusion

The three years since the onset of the COVID-19 pandemic has seen profound societal change in how individuals live, work and interact with each other. However, the impact of these changes on patterns of weight gain have not been well characterised. We are, to our knowledge, the first to use a nationally representative population database^16^ to describe patterns of weight gain during the pandemic. We report novel findings from our analyses of BMI data in the EHRs of over 17 million adults living in England. At population level there was a modest increase in the median BMI between the prepandemic year and the year beginning March 2021. At individual-level we showed that, amongst adults with BMI values recorded in the EHR, women, younger adults and those living in the most socioeconomically deprived areas were at greatest estimated risk of unhealthy patterns of weight gain, namely rapid weight gain during the pandemic and an extreme acceleration in rate of weight gain between the prepandemic and pandemic periods. Individuals with a history of LTCs were also at greatest risk of unhealthy weight gain, and people with mental health conditions, including depression, SMI and learning difficulties, were at disproportionately greater risk than those with physical health conditions such as hypertension.

Previous research in this area has been limited to small voluntary surveys,^10^ or routine health data from non-representative populations.^11–14^ By using a large-scale, nationally representative and contemporaneous EHR dataset, we have been able to robustly reproduce prior observations that women,^11,12^ young adults,^11,12^ those living in deprivation^13^ and those with LTCs^13,14^ may be the most affected by unhealthy patterns of pandemic weight gain and demonstrate this at population scale. Our findings suggest the increased risk of weight gain seen amongst young adults prepandemic^15^ was exacerbated during the pandemic. The increased risk seen amongst women may reflect gender disparities in the impact of the pandemic on social factors such as employment loss and caring responsibilities, with a consequent influence on health behaviours.^18^ The increased risk amongst those living in areas of greater deprivation reflects prepandemic trends and is likely to have multifactorial aetiology including food poverty, reduced opportunity for physical activity and an increased burden of physical and mental health conditions.^19^ In our analysis, instead of controlling for LTCs as prior studies have done,^1,15^ we characterised the associations between patterns of weight gain and clinical and mental health characteristics. This approach uncovered important inequality between people with prior mental health versus physical health conditions, with the former having a greater risk of unhealthy patterns of weight gain. These differences may reflect associations between disordered eating, reduced physical activity and poor mental health exacerbated by the pandemic.^20,21^ Our finding highlights the need for health services to ensure parity of esteem^22^ between physical and mental health conditions when prioritising groups to be supported by weight loss interventions.

The key strengths of this study are the quality, scale and representativeness of the health record data used.^16^ OpenSAFELY-TPP provides access to the primary care records of roughly 40% of the population.^16^ In England, primary care hosts and records data from 90% of all patient consultations in the National Health Service, with particular responsibility for preventative care and routine management of LTCs.^23^ Therefore, our study has been able to make robust observations that have immediate relevance to the delivery of health care in the post-pandemic period. These findings could be rapidly implemented in the routine clinical care that has generated them, for example through the practice reorganisation or redistribution of care to target those at most need.

We recognise the limitations of our work. We restricted our analyses to individuals registered with a GP practice for at least one year to minimise the impact of incomplete data. However, this may have introduced survival bias, e.g. if individuals with the most unhealthy patterns of weight gain were more likely to die during the pandemic. BMI is recorded in EHRs when clinically indicated, this may be systematically as part of the routine monitoring of LTCs such as T2D or opportunistically when relevant to the clinical consultation. Despite this risk of information bias, prepandemic EHRs produced BMI trajectory estimates comparable to representative national surveys.^24^ However, we report changes in BMI recording activity that may have introduced new bias, with a decline in BMI recording after the onset of the pandemic which had only partly recovered by the year beginning March 2021, reflecting wider patterns of primary care activity during the pandemic.^25^ Notably, the greatest recovery in BMI recording was amongst individuals with LTCs requiring annual BMI recording, e.g. T2D. This leads to a higher prevalence of LTCs amongst individuals contributing pandemic BMI data (Table 1) and the possibility that the protective effect of T2D against weight gain is in fact due to collider bias.^26^ Alternatively this may be a true effect arising from the awareness that COVID-19 infection confers greater risk to people with obesity and LTCs such as T2D, leading to more proactive and intensive weight management in primary care, or adoption of healthy behaviours. The observed protective effect of T1D against extreme acceleration may also reflect bias, however there is evidence of improved blood glucose control amongst people living with T1D during the pandemic.^27^ Pandemic-associated changes in clinical activity, such as BMI self-reporting during remote consultations, may have introduced biases that are harder to characterise.^28^ Further research, including representative population health surveys, would be required to investigate this further.

Another limitation relates to missing data. We have undertaken a complete case analysis, removing individuals with missing ethnicity and IMD data. This approach precludes generalisability of findings to those missing data in these fields, e.g. those missing IMD data due to unstable accommodation, but gives robust evidence in everybody else. Multiple imputation models would not overcome this limitation, as the Missing at Random assumption is unlikely to hold.^29^ Some prepandemic analyses of BMI trajectories have used imputation, adjusting estimates with population-survey data to account for the MNAR assumption.^15^ We could not replicate this approach due to the suspension of such surveys during the pandemic.

In conclusion, our study is the largest and most representative analysis of weight changes associated with the pandemic to date. We identify that women, young adults, those living in the most deprivation, as well as those with mental health conditions were at increased risk of unhealthy patterns of weight gain during the pandemic. We demonstrate the value of routinely collected data to understand the impact of a pandemic at population scale and identify health inequalities and areas of focus. Overall, our findings identified clear targets for health services and policy makers to target with preventative interventions to mitigate the lasting and inequitable impact of the pandemic. Future work is needed to continue the monitoring of these trends to understand whether these effects are lasting or even exacerbated.

## Data Availability

All data were linked, stored and analysed securely within the OpenSAFELY-TPP platform, https://opensafely.org/, containing pseudonymised data on approximately 40% of the English population, including coded diagnoses, medications and physiological parameters. No free text data are included. Detailed pseudonymised patient data is potentially re-identifiable and therefore not shared. All code for data management and analysis, as well as codelists is shared openly for review and re-use under MIT open license, available at https://github.com/opensafely/BMI-and-Metabolic-Markers.

## Role of Funding Source

This study was undertaken by MS as part of her National Institute of Health Care Research (NIHR) funded academic clinical fellowship in primary care. There was no other direct funding for this analysis.

## Declaration of Interests

MS salary costs have been supported through a National Institute for Health and Care Research (NIHR) funded academic clinical fellowship in primary care and NIHR grant funding (NIHR AI-MULTIPLY Consortium NIHR203982). RYP is supported by the EPSRC Centre for Doctoral Training in Health Data Science (EP/S02428X/1). RYP was previously employed as a data scientist for the Bennet Institute which is funded by grants from the Bennett Foundation, Wellcome Trust, NIHR Oxford Biomedical Research Centre, NIHR Applied Research Collaboration Oxford and Thames Valley, Mohn-Westlake Foundation. SVE is funded by a Diabetes UK Sir George Alberti research training fellowship (grant number: 17/0005588). FE salary cost is supported by MRC (MR/S027297/1) “Multimorbidity, clusters, trajectories and genetic risk in British south Asians, 2020-2023”. DS is funded by the NIHR (NIHR203982). AM is a senior clinical researcher at the University of Oxford in the Bennett Institute, which is funded by grants from the Bennett Foundation, Wellcome Trust, NIHR Oxford Biomedical Research Centre, NIHR Applied Research Collaboration Oxford and Thames Valley, Mohn-Westlake Foundation. AM has consulted for https://inductionhealthcare.com/. AM is a member of the RCGP health informatics group and the NHS Digital GP data Professional Advisory Group that advises on access to GP Data for Pandemic Planning and Research (GDPPR); payment direct to me for the GDPPR role. RM is supported by Barts Charity (MGU0504). JV is National Clinical Director for Diabetes & Obesity at NHS England. BMK is also employed by NHS England. KK is supported by the National Institute for Health Research (NIHR) Applied Research Collaboration East Midlands (ARC EM) and the NIHR Leicester Biomedical Research Centre (BRC). KK has acted as a consultant, speaker or received grants for investigator-initiated studies for Astra Zeneca, Bayer, Novartis, Novo Nordisk, Sanofi-Aventis, Lilly and Merck Sharp & Dohme, Boehringer Ingelheim, Oramed Pharmaceuticals, Roche and Applied Therapeutics. SF has received grants from the NIHR (NIHR 31672, NIHR 202635) and MRC (MR/W014416/1, MR/V004905/1, MR/S027297/1). SF, RM, CM are part of the Genes & Health programme, which is part-funded (including salary contributions) by a Life Sciences Consortium comprising Astra Zeneca PLC, Bristol-Myers Squibb Company, GlaxoSmithKline Research and Development Limited, Maze Therapeutics Inc, Merck Sharp & Dohme LLC, Novo Nordisk A/S, Pfizer Inc, Takeda Development Centre Americas Inc.

This research used data assets made available as part of the Data and Connectivity National Core Study, led by Health Data Research UK in partnership with the Office for National Statistics and funded by UK Research and Innovation (grant ref MC_PC_20058). In addition, the OpenSAFELY Platform is supported by grants from the Wellcome Trust (222097/Z/20/Z); MRC (MR/V015757/1, MC_PC-20059, MR/W016729/1); NIHR (NIHR135559, COV-LT2-0073), and Health Data Research UK (HDRUK2021.000, 2021.0157).

## Data Sharing

Access to the underlying identifiable and potentially re-identifiable pseudonymised electronic health record data is tightly governed by various legislative and regulatory frameworks, and restricted by best practice. The data in OpenSAFELY-TPP is drawn from General Practice data across England where TPP is the data processor.

TPP developers initiate an automated process to create pseudonymised records in the core OpenSAFELY database, which are copies of key structured data tables in the identifiable records. These pseudonymised records are linked onto key external data resources that have also been pseudonymised via SHA-512 one-way hashing of NHS numbers using a shared salt. Bennett Institute for Applied Data Science developers and PIs holding contracts with NHS England have access to the OpenSAFELY pseudonymised data tables as needed to develop the OpenSAFELY tools.

These tools in turn enable researchers with OpenSAFELY Data Access Agreements to write and execute code for data management and data analysis without direct access to the underlying raw pseudonymised patient data, and to review the outputs of this code. All code for the full data management pipeline—from raw data to completed results for this analysis—and for the OpenSAFELY platform as a whole is available for review at https://github.com/OpenSAFELY. The data management and analysis code for this paper was led by MS and RYP and is available for scientific review and re-use under MIT open licence. https://github.com/opensafely/BMI-and-Metabolic-Markers.

## Acknowledgements

We are very grateful for all the support received from the TPP Technical Operations team throughout this work, and for generous assistance from the information governance and database teams at NHS England and the NHS England Transformation Directorate.

## Authors Contribution Statement

All authors contributed to this manuscript. MS, RYP, RM, CEM, BMK, KK, JV and SF contributed to the conceptualisation of the study. MS, RYP, CEM, SB, AM, JM, ID, PI, WJH, RR, BMK and SF were involved in project administration underpinning this analysis. CEM, SB, AM, JM, ID, PI, WJH, and BMK contributed to the development and maintenance of the OpenSAFELY-TPP platform. MS, RYP, CEM, SB, AM, ID, PI and WJH were involved in curation of the data used in the analysis. MS, RYP, SVE, FE, DS, RM and SF contributed to the development and design of the statistical methodology used in these analyses. MS conducted the formal analysis including the application of statistical techniques with technical support from RYP and further support and supervision from DS, SVE, RM and SF. RYP, SVE, WJH, KK, RM, JV, BMK and SF contributed to supervision of different parts of the analysis including support to use the platform, support in the analyses and interpretation of the study findings. MS wrote the original draft. All authors contributed to further review and editing of the draft.

## Notes

### Author Declarations

The London School of Hygiene & Tropical Medicine Ethics Board (reference 26536)gave ethical approval for this work

### Summary of Updates

No changes have been made to the revised file.

