## Supplementary Files for "Trends in weight gain recorded in English primary care before and during the Coronavirus-19 pandemic: an observational cohort study using the OpenSAFELY platform"

#### Supplementary File

### Appendix 1. Calculation of δ-prepandemic, δ-pandemic and δ-change

BMIs were classified into time periods: period-1: March 2015 - February 2018; period-2: March 2018 - February 2020; and period-3 March 2020 - February 2022. For patients with more than one BMI measure from each time period, a random BMI measure was selected from each time period. Where data were available, BMI data from period 1 and period 2 were used to calculate δ-prepandemic (e.g. dummy patient 1), while BMI data from period 2 and period 3 were used to calculate δ-pandemic (e.g dummy patient 1 and dummy patient 2). Rate of BMI change/year was calculated between these time points assuming a linear trend (Katsoulis, Lai, et al. 2021).

To assess the impact of the pandemic in rate of weight gain, we calculated delta change as the change in rate of weight gain between the prepandemic and pandemic periods (δ-change = δ-pandemic - δ-prepandemic). Patients therefore required both a δ-prepandemic and δ-pandemic value to contribute to the δ-change analysis (e.g. dummy patient 1). Patients with either a δ-prepandemic or δ-pandemic, but not both, contributed to the prepandemic or pandemic BMI trajectory analysis respectively, but not to the δ-change analysis (e.g. dummy patient 2). Patients without BMI data from period 2 were unable to contribute to any of these analyses (e.g. dummy patient 3).

**Supplementary Figure 1. Demonstration of calculation of δ-prepandemic, δ-pandemic and δ-change using dummy patient data**

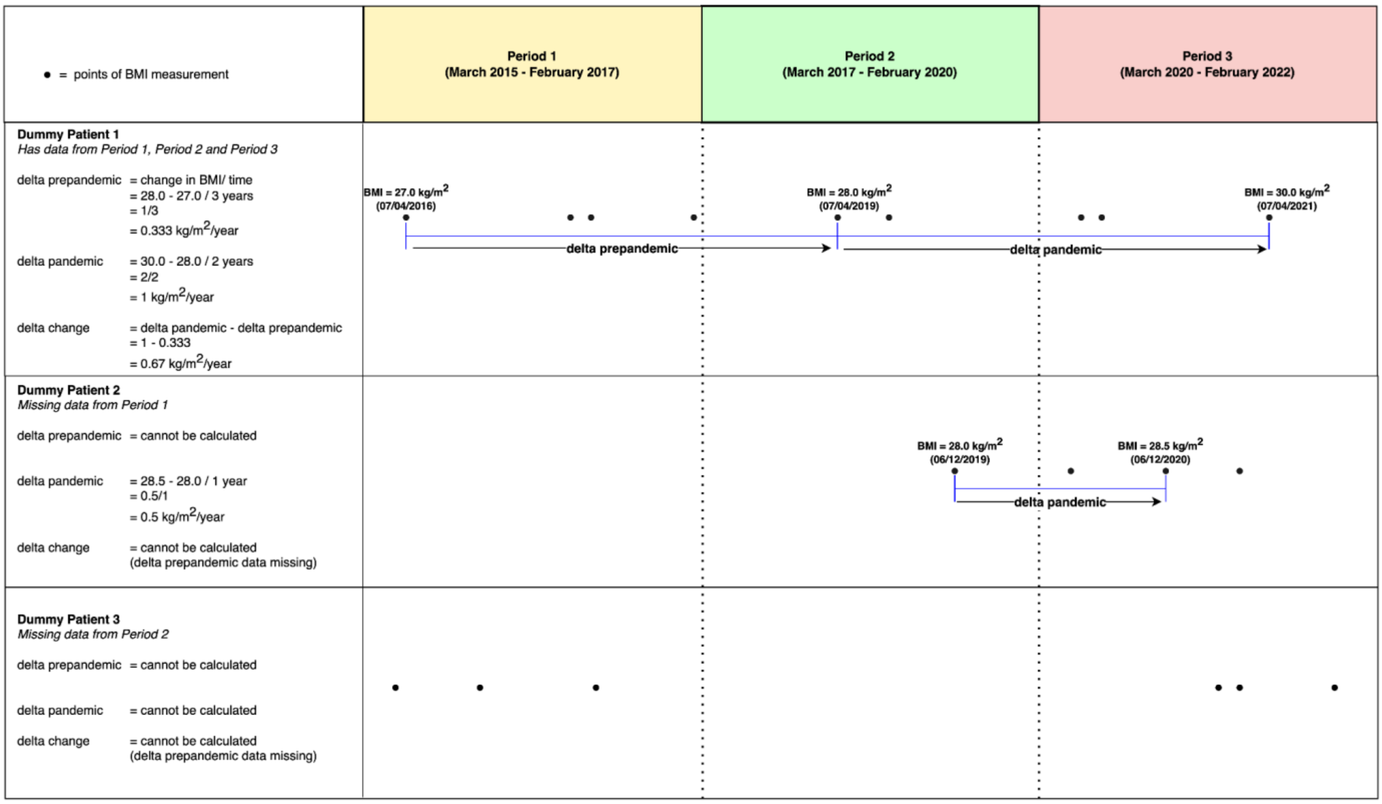

#### Appendix 2. Defining a population of extreme accelerators (distribution of δ-change)

We plotted the distribution of δ-change in the 2,768,695 individuals contributing to the analysis of change in rate of weight gain after the onset of the pandemic. Supplementary Figure 1 demonstrates the deciles of δ-change. We identified individuals with a δ-change >= 1.84 kg/m$^2$/year as the ten percent of the population experiencing the most extreme acceleration in their rate of weight gain during the pandemic and explored the distribution of extreme accelerators in different subgroups of the study population.

As extreme accelerators were defined as the top-decile of estimated δ-change in the total study population, clinical and sociodemographic subgroups in which over ten percent of individuals were extreme accelerators had an increased absolute estimated risk of extreme acceleration, conversely subgroups in which less than ten percent were extreme accelerators had a reduced absolute estimated risk.

**Supplementary Figure 2. Cumulative distribution of change in rate of weight gain (δ-change) amongst adults living in England before and after the onset of the COVID-19 pandemic.**

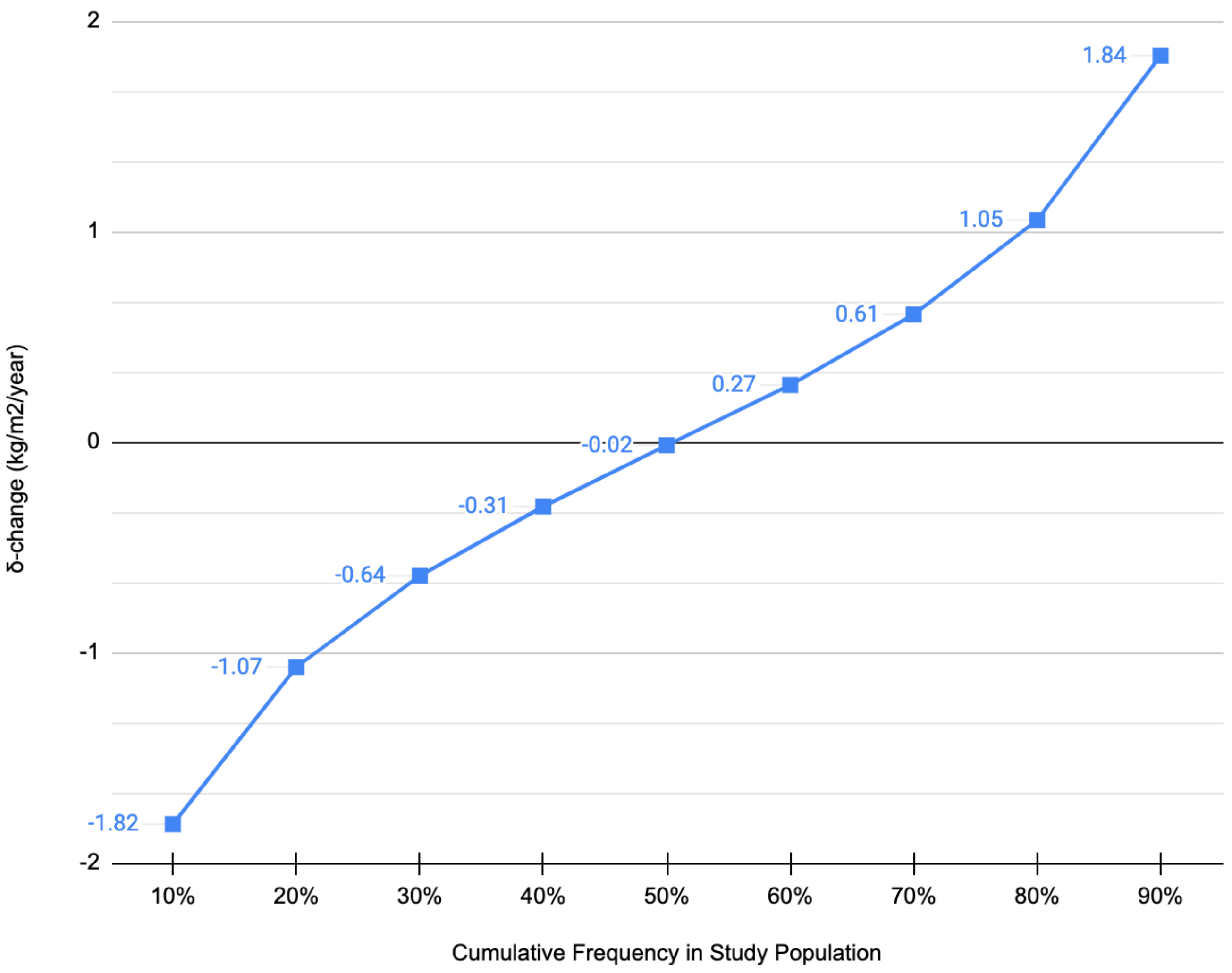

#### Appendix 3. Information Governance Statement

NHS England is the data controller for OpenSAFELY-TPP; TPP is the data processor; all study authors using OpenSAFELY have the approval of NHS England. This implementation of OpenSAFELY is hosted within the TPP environment which is accredited to the ISO 27001 information security standard and is NHS IG Toolkit compliant. Patient data has been pseudonymised for analysis and linkage using industry standard cryptographic hashing techniques; all pseudonymised datasets transmitted for linkage onto OpenSAFELY are encrypted; access to the platform is via a virtual private network (VPN) connection, restricted to a small group of researchers; the researchers hold contracts with NHS England and only access the platform to initiate database queries and statistical models; all database activity is logged; only aggregate statistical outputs leave the platform environment following best practice for anonymisation of results such as statistical disclosure control for low cell counts. The OpenSAFELY research platform adheres to the obligations of the UK General Data Protection Regulation (GDPR) and the Data Protection Act 2018. In March 2020, the Secretary of State for Health and Social Care used powers under the UK Health Service (Control of Patient Information) Regulations 2002 (COPI) to require organisations to process confidential patient information for the purposes of protecting public health, providing healthcare services to the public and monitoring and managing the COVID-19 outbreak and incidents of exposure; this sets aside the requirement for patient consent. This was extended in November 2022 for the NHS England OpenSAFELY COVID-19 research platform. In some cases of data sharing, the common law duty of confidence is met using, for example, patient consent or support from the Health Research Authority Confidentiality Advisory Group. Taken together, these provide the legal bases to link patient datasets on the OpenSAFELY platform. GP practices, from which the primary care data are obtained, are required to share relevant health information to support the public health response to the pandemic, and have been informed of the OpenSAFELY analytics platform. This study was supported by Dr Jonathan Valabhji, clinical director for diabetes and obesity at NHS England, as senior sponsor.

1. Data Security and Protection Toolkit - NHS Digital. NHS Digital. **https://digital.nhs.uk/data-and-information/looking-after-information/data-security-and-information-governance/data-security-and-protection-toolkit** (accessed 30 Apr 2020). ↩
2. ISB1523: Anonymisation Standard for Publishing Health and Social Care Data - NHS Digital. NHS Digital. **https://digital.nhs.uk/data-and-information/information-standards/information-standards-and-data-collections-including-extractions/publications-and-notifications/standards-and-collections/isb1523-anonymisation-standard-for-publishing-health-and-social-care-data** (accessed 30 Apr 2020). ↩
3. Secretary of State for Health and Social Care - UK Government. Coronavirus (COVID-19): notification to organisations to share information. 2020. **https://web.archive.org/web/20200421171727/https://www.gov.uk/government/publications/covid-19-notification-to-gps-and-nhs-england-to-share-information** ↩
4. Secretary of State for Health and Social Care - UK Government. Coronavirus (COVID-19): notification to organisations to share information. 2022. **https://www.gov.uk/government/publications/coronavirus-covid-19-notification-to-organisations-to-share-information/coronavirus-covid-19-notice-under-regulation-34-of-the-health-service-control-of-patient-information-regulations-2002** ↩
5. Confidentiality Advisory Group. Health Research Authority. **https://www.hra.nhs.uk/about-us/committees-and-services/confidentiality-advisory-group/** ↩

### Supplementary Table 1: Proportion of adults living in England with a BMI value recorded in their routine health care record

| Supplementary Table 1: Proportion of adults living in England with at least one Body Mass Index (BMI) value recorded in their routine electronic health care records per year before and after the onset of the Coronavirus-19 (COVID-19) pandemic | | | | | | |
|---|---|---|---|---|---|---|
| | **Year Beginning March 2021** | | **Year Beginning March 2020** | | **Prepandemic Year Beginning March 2019** | |
| **Population Subgroup** | **Study Populatio N (%)** | **BMI recorded % (95% CI)** | **Study Population N (%)** | **BMI recorded % (95% CI)** | **Study Population N (%)** | **BMI recorded % (95% CI)** |
| Total | 17,742,365 | 24·99 (24·97, 25·01) | 17,037,385 | 18·69 (18·67, 18·70) | 16,301,350 | 31·30 (31·28, 31·33) |
| Age Group (years) | | | | | | |
| 18-29 | 3,062,705 (17·3) | 14·90 (14·86, 14·94) | 2,956,395 (17·4) | 13·94 (13·90, 13·98) | 2,822,850 (17·3) | 20·66 (20·61, 20·71) |
| 30-39 | 3,138,645 (17·7) | 15·03 (14·99, 15·07) | 3,018,435 (17·7) | 12·81 (12·77, 12·85) | 2,889,440 (17·7) | 20·11 (20·06, 20·15) |
| 40-49 | 2,953,105 (16·6) | 20·08 (20·03, 20·12) | 2,893,890 (17·0) | 14·46 (14·42, 14·50) | 2,826,860 (17·3) | 26·04 (25·99, 26·09) |
| 50-59 | 3,176,680 (17·9) | 26·00 (25·95, 26·04) | 3,082,985 (18·1) | 18·16 (18·12, 18·21) | 2,987,265 (18·3) | 32·14 (32·08, 32·19) |
| 60-69 | 2,495,010 (14·1) | 33·73 (33·67, 33·78) | 2,386,965 (14·0) | 24·28 (24·23, 24·34) | 2,302,715 (14·1) | 42·63 (42·56, 42·69) |
| 70-79 | 1,997,575 (11·3) | 41·92 (41·85, 41·99) | 1,878,465 (11·0) | 29·98 (29·91, 30·04) | 1,756,870 (10·8) | 50·45 (50·37, 50·52) |
| 80-89 | 918,640 (5·2) | 44·41 (44·31, 44·52) | 820,250 (4·8) | 32·12 (32·02, 32·22) | 715,345 (4·4) | 52·36 (52·25, 52·48) |
| Sex | | | | | | |
| Female | 8,883,720 (50·1) | 29·25 (29·22, 29·28) | 8,538,025 (50·1) | 21·98 (21·95, 22·01) | 8,180,085 (50·2) | 36·04 (36·01, 36·07) |
| Male | 8,858,645 (49·9) | 20·71 (20·69, 20·74) | 8,499,360 (49·9) | 15·37 (15·35, 15·40) | 8,121,265 (49·8) | 26·53 (26·50, 26·56) |
| Patient IMD | | | | | | |
| 1 (most deprived) | 3,356,720 (18·9) | 25·95 (25·90, 26·00) | 3,240,870 (19·0) | 19·72 (19·67, 19·76) | 3,102,370 (48·4) | 32·67 (32·62, 32·72) |
| 5 (least deprived) | 3,240,120 (18·3) | 24·05 (24·01, 24·10) | 3,133,315 (18·4) | 17·67 (17·62, 17·71) | 3,029,530 (47·3) | 29·93 (29·88, 29·98) |
| Missing | 431,235 (2·4) | 25·16 (25·03, 25·29) | 345,395 (2·0) | 19·16 (19·02, 19·29) | 273,985 (4·3) | 32·62 (32·45, 32·80) |
| Ethnicity | | | | | | |
| White British | 12,299,145 (76·1) | 28·10 (28·07, 28·12) | 11,944,365 (76·8 | 20·83 (20·81, 20·86) | 11,590,555 (77·8) | 34·72 (34·70, 34·75) |
| White Irish | 93,035 (0·6) | 26·36 (26·07, 26·64) | 88,600 (0·6) | 19·76 (19·50, 20·03) | 84,305 (0·6) | 32·96 (32·64, 33·28) |
| Other White | 1,540,215 (9·5) | 16·57 (16·51, 16·63) | 1,437,855 (9·2) | 12·89 (12·83, 12·94) | 1,316,480 (8·8) | 22·30 (22·23, 22·37) |
| Indian | 501,025 (3·1) | 25·99 (25·87, 26·12) | 469,700 (3·0) | 19·54 (19·43, 19·66) | 434,445 (2·9) | 32·17 (32·03, 32·30) |
| Pakistani | 358,585 (2·2) | 27·69 (27·55, 27·84) | 340,615 (2·2) | 20·85 (20·72, 20·99) | 319,705 (2·1) | 34·89 (34·72, 35·05) |
| Bangladeshi | 83,300 (0·5) | 28·56 (28·26, 28·87) | 78,085 (0·5) | 22·24 (21·95, 22·53) | 72,560 (0·5) | 34·94 (34·59, 35·28) |
| Chinese | 115,825 (0·7) | 11·17 (10·99, 11·35) | 106,645 (0·7) | 9·00 (8·83, 9·17) | 93,050 (0·6) | 17·08 (16·83, 17·32) |
| Other Asian | 264,080 (1·6) | 23·67 (23·50, 23·83) | 245,040 (1·6) | 18·29 (18·14, 18·44) | 223,795 (1·5) | 29·35 (29·16, 29·54) |
| Black Caribbean | 95,140 (0·6) | 31·09 (30·80, 31·39) | 91,260 (0·6) | 23·24 (22·96, 23·51) | 86,680 (0·6) | 37·62 (37·30, 37·95) |
| Black African | 219,880 (1·4) | 21·16 (20·99, 21·33) | 202,735 (1·3) | 16·40 (16·24, 16·56) | 183,060 (1·2) | 26·97 (26·77, 27·17) |
| Other Black | 89,040 (0·6) | 23·95 (23·67, 24·23) | 82,465 (0·5) | 17·63 (17·37, 17·89) | 75,545 (0·5) | 30·26 (29·94, 30·59) |
| White & Black Caribbean | 49,930 (0·3) | 23·42 (23·04, 23·79) | 46,580 (0·3) | 19·19 (18·83, 19·55) | 43,290 (0·3) | 30·22 (29·78, 30·65) |
| White & Black African | 39,065 (0·2) | 19·80 (19·41, 20·20) | 35,720 (0·2) | 15·90 (15·52, 16·28) | 32,370 (0·2) | 25·91 (25·43, 26·38) |
| White & Asian | 42,740 (0·3) | 19·99 (19·62, 20·37) | 39,050 (0·3) | 16·07 (15·70, 16·43) | 35,450 (0·2) | 26·14 (25·68, 26·59) |
| Other Mixed | 82,940 (0·5) | 18·76 (18·50, 19·03) | 76,315 (0·5) | 15·39 (15·13, 15·64) | 69,245 (0·5) | 24·68 (24·36, 25·01) |
| Other | 286,925 (1·8) | 18·84 (18·70, 18·98) | 261,295 (1·7) | 15·25 (15·12, 15·39) | 234,955 (1·6) | 24·55 (24·38, 24·73) |
| Missing | 1,581,510 (8·9) | 11·04 (10·99, 11·09) | 1,491,055 (8·8) | 7·80 (7·76, 7·84) | 1,405,850 (8·6) | 13·38 (13·33, 13·44) |
| Long Term Condition | | | | | | |
| Hypertension | 3,558,405 (20·1) | 49·22 (49·17, 49·27) | 3,430,595 (20·1) | 36·07 (36·02, 36·12) | 3,230,230 (19·8) | 56·92 (56·86, 56·97) |
| Type 2 Diabetes | 1,231,455 (6·9) | 72·18 (72·10, 72·26) | 1,151,500 (6·8) | 58·21 (58·12, 58·30) | 1,051,605 (6·5) | 82·54 (82·46, 82·61) |
| Type 1 Diabetes | 93,505 (0·5) | 59·59 (59·28, 59·90) | 87,905 (0·5) | 47·25 (46·92, 47·58) | 83,455 (0·5) | 71·25 (70·94, 71·56) |
| Learning Disability | 105,855 (0·6) | 62·03 (61·74, 62·32) | 99,295 (0·6) | 51·87 (51·56, 52·18) | 93,420 (0·6) | 65·65 (65·34, 65·95) |
| Depression | 3,592,080 (20·2) | 31·85 (31·80, 31·89) | 3,415,085 (20·0) | 23·70 (23·65, 23·74) | 3,197,730 (19·6) | 38·66 (38·60, 38·71) |
| Dementia | 134,080 (0·8) | 45·57 (45·31, 45·84) | 114,135 (0·7) | 35·50 (35·22, 35·78) | 90,895 (0·6) | 53·45 (53·13, 53·78) |
| Serious Mental Illness * | 195,450 (1·1) | 60·56 (60·34, 60·77) | 182,630 (1·1) | 48·65 (48·42, 48·88) | 169,995 (1·0) | 68·38 (68·16, 68·60) |
| COPD** | 531,475 (3·0) | 49·49 (49·36, 49·63) | 506,180 (3·0) | 36·04 (35·91, 36·17) | 458,960 (2·8) | 67·25 (67·12, 67·39) |
| Asthma | 2,930,645 (16·5) | 29·82 (29·76, 29·87) | 2,801,825 (16·4) | 22·74 (22·69, 22·79) | 2,660,330 (16·3) | 39·20 (39·14, 39·25) |
| Cardiovascular Disease | 1,087,320 (6·1) | 53·31 (53·22, 53·40) | 1,021,155 (6·0) | 39·98 (39·89, 40·08) | 940,115 (5·8) | 62·69 (62·59, 62·78) |

| | | | | | | |
|---|---|---|---|---|---|---|
| Stroke and TIA | 465,645 (2·6) | 49·49 (49·35, 49·64) | 428,365 (2·5) | 36·81 (36·66, 36·95) | 387,020 (2·4) | 58·56 (58·41, 58·72) |
| Cancer | 905,475 (5·1) | 38·54 (38·44, 38·64) | 839,895 (4·9) | 27·80 (27·70, 27·89) | 766,395 (4·7) | 45·52 (45·41, 45·63) |

* Serious Mental Illness includes Psychosis and Bipolar Disorder
** Chronic Obstructive Pulmonary Disease (COPD)

#### Supplementary Table 2: Average rate of weight gain amongst adults living in England before and after the onset of the COVID-19 pandemic

| Supplementary Table 2: Rate of weight gain in $kg/m^2$/year amongst adults living in England before and after the onset of the COVID-19 pandemic | | | | |
|---|---|---|---|---|
| | **Pandemic rate of weight gain** | | **Prepandemic rate of weight gain** | |
| | **N (%)** | **Mean (SD)** | **N (%)** | **Mean (SD)** |
| Total Population | 3,214,155 | 0·06 (1·20) | 3,966,495 | 0·08 (1·05) |
| **Sex** | | | | |
| Female | 1,898,510 (59·1) | 0·13 (1·27) | 2,381,345 (60·0) | 0·12 (1·12) |
| Male | 1,315,645 (40·9) | -0·03 (1·08) | 1,585,155 (40·0) | 0·02 (0·94) |
| Age Group | | | | |
| 18-29 | 267,605 (8·3) | 0·40 (1·49) | 274,540 (6·9) | 0·45 (1·36) |
| 30-39 | 341,870 (10·6) | 0·30 (1·40) | 466,285 (11·8) | 0·28 (1·23) |
| 40-49 | 415,805 (12·9) | 0·17 (1·27) | 545,760 (13·8) | 0·17 (1·12) |
| 50-59 | 603,670 (18·8) | 0·07 (1·19) | 757,830 (19·1) | 0·08 (1·05) |
| 60-69 | 642,120 (20·0) | -0·00 (1·09) | 771,890 (19·5) | -0·00 (0·96) |
| 70-79 | 635,175 (19·8) | -0·08 (1·01) | 757,405 (19·1) | -0·06 (0·87) |
| 80-89 | 307,910 (9·6) | -0·21 (1·04) | 392,785 (9·9) | -0·15 (0·84) |
| **Ethnicity** | | | | |
| White British | 2,631,235 (81·9) | 0·07 (1·21) | 3,275,985 (82·6) | 0·07 (1·06) |
| White Irish | 17,590 (0·5) | -0·01 (1·18) | 21,500 (0·5) | 0·02 (1·02) |
| Other White | 172,260 (5·4) | 0·10 (1·24) | 211,145 (5·3) | 0·12 (1·08) |
| Indian | 99,750 (3·1) | -0·03 (1·02) | 115,585 (2·9) | 0·03 (0·92) |
| Pakistani | 80,400 (2·5) | 0·05 (1·09) | 99,045 (2·5) | 0·10 (0·98) |
| Bangladeshi | 18,965 (0·6) | 0·01 (1·03) | 21,230 (0·5) | 0·10 (0·94) |
| Chinese | 8,245 (0·3) | -0·04 (0·91) | 10,185 (0·3) | 0·04 (0·80) |
| Other Asian | 46,590 (1·4) | 0·02 (1·04) | 51,865 (1·3) | 0·08 (0·94) |
| Black African | 32,875 (1·0) | 0·14 (1·21) | 37,745 (1·0) | 0·13 (1·07) |
| Black Caribbean | 23,050 (0·7) | 0·06 (1·19) | 27,170 (0·7) | 0·03 (1·03) |
| Other Black | 15,020 (0·5) | 0·12 (1·23) | 17,295 (0·4) | 0·10 (1·10) |
| Mixed White/Black Caribbean | 8,840 (0·3) | 0·16 (1·35) | 10,610 (0·3) | 0·15 (1·18) |
| Mixed White/ Black African | 5,460 (0·2) | 0·15 (1·27) | 6,400 (0·2) | 0·15 (1·07) |
| Mixed White/Asian | 5,945 (0·2) | 0·08 (1·19) | 7,055 (0·2) | 0·13 (1·08) |
| Other Mixed | 10,795 (0·3) | 0·14 (1·27) | 12,525 (0·3) | 0·16 (1·13) |
| Other | 37,130 (1·2) | 0·06 (1·17) | 41,155 (1·0) | 0·10 (1·06) |
| **Patient IMD** | | | | |
| 1 (most deprived) | 686,100 (21·3) | 0·12 (1·29) | 836,480 (21·1) | 0·11 (1·14) |
| 2 | 659,115 (20·5) | 0·08 (1·23) | 804,470 (20·3) | 0·08 (1·08) |
| 3 | 692,270 (21·5) | 0·05 (1·18) | 849,790 (21·4) | 0·07 (1·04) |
| 4 | 627,180 (19·5) | 0·04 (1·15) | 783,690 (19·8) | 0·06 (1·01) |
| 5 (least deprived) | 549,495 (17·1) | 0·01 (1·11) | 692,065 (17·4) | 0·06 (0·97) |
| **Long Term Condition** | | | | |
| Hypertension | | | | |
| Absent | 1,778,085 (55·3) | 0·15 (1·26) | 2,329,335 (58·7) | 0·15 (1·11) |
| Present | 1,436,070 (44·7) | -0·04 (1·11) | 1,637,165 (41·3) | -0·03 (0·96) |
| Type 1 Diabetes | | | | |
| Absent | 3,162,905 (98·4) | 0·06 (1·20) | 3,910,690 (98·6) | 0·08 (1·05) |
| Present | 51,250 (1·6) | 0·18 (1·11) | 55,810 (1·4) | 0·13 (0·96) |
| Type 2 Diabetes | | | | |
| Absent | 2,420,385 (75·3) | 0·14 (1·22) | 3,146,110 (79·3) | 0·13 (1·06) |
| Present | 793,770 (24·7) | -0·17 (1·12) | 820,390 (20·7) | -0·13 (0·99) |
| Cardiovascular Disease | | | | |
| Absent | 2,726,240 (84·8) | 0·09 (1·21) | 3,406,465 (85·9) | 0·10 (1·07) |
| Present | 487,915 (15·2) | -0·08 (1·11) | 560,030 (14·1) | -0·05 (0·94) |
| Learning Difficulties | | | | |
| Absent | 3,158,405 (98·3) | 0·06 (1·20) | 3,911,915 (98·6) | 0·08 (1·05) |
| Present | 55,750 (1·7) | 0·15 (1·41) | 54,580 (1·4) | 0·15 (1·23) |
| Depression | | | | |
| Absent | 2,326,915 (72·4) | 0·04 (1·14) | 2,860,955 (72·1) | 0·06 (1·00) |
| Present | 887,240 (27·6) | 0·12 (1·34) | 1,105,540 (27·9) | 0·12 (1·17) |
| Dementia | | | | |
| Absent | 3,166,930 (98·5) | 0·07 (1·20) | 3,907,830 (98·5) | 0·08 (1·05) |
| Present | 47,225 (1·5) | -0·23 (1·39) | 58,670 (1·5) | -0·15 (1·09) |
| Serious Mental Illness (Psychosis and Bipolar Disorder) | | | | |
| Absent | 3,112,755 (96·8) | 0·06 (1·19) | 3,866,475 (97·5) | 0·08 (1·05) |
| Present | 101,395 (3·2) | 0·13 (1·47) | 100,025 (2·5) | 0·11 (1·26) |
| Asthma | | | | |
| Absent | 2,504,425 (77·9) | 0·06 (1·17) | 3,026,570 (76·3) | 0·07 (1·04) |
| Present | 709,730 (22·1) | 0·09 (1·29) | 939,925 (23·7) | 0·10 (1·11) |
| Chronic Obstructive Pulmonary Disease | | | | |
| Absent | 2,986,215 (92·9) | 0·07 (1·20) | 3,667,400 (92·5) | 0·08 (1·06) |
| Present | 227,935 (7·1) | -0·05 (1·21) | 299,095 (7·5) | 0·01 (1·01) |
| Stroke and TIA | | | | |
| Absent | 3,026,575 (94·2) | 0·07 (1·20) | 3,747,140 (94·5) | 0·08 (1·06) |
| Present | 187,580 (5·8) | -0·09 (1·14) | 219,360 (5·5) | -0·06 (0·96) |

### Supplementary Table 3: Age stratified associations with risk of extreme acceleration in rate of weight gain

**Supplementary Table 3: Age stratified analysis of associations between sociodemographic and clinical characteristics and risk of extreme acceleration in rate of weight gain (δ-change >= 1·84 $kg/m^2$/year) during the pandemic.**

| | Age Group: 18 – 39 Years | | | Age Group: 40 - 59 Years | | | Age Group: 60 - 79 Years | | |
|---|---|---|---|---|---|---|---|---|---|
| | N (%) | AR(%) | aOR (95% CI) | N (%) | AR (%) | aOR (95% CI) | N (%) | AR (%) | aOR (95% CI) |
| **Sex** | | | | | | | | | |
| F | 361,125 (87·00) | 14·77 | 1 | 478,495 (60·59) | 12·28 | 1 | 589,805 (48·23) | 10·03 | 1 |
| M | 53,950 (13·00) | 13·60 | 0·91 (0·89 to 0·94) | 311,205 (39·41) | 9·55 | 0·76 (0·75 to 0·77) | 633,150 (51·77) | 6·94 | 0·67 (0·66 to 0·68) |
| **Ethnicity** | | | | | | | | | |
| White | 365,300 (88·01) | 14·84 | 1 | 650,425 (82·36) | 11·81 | 1 | 1,116,720 (91·31) | 8·60 | 1 |
| Black | 7,530 (1·81) | 15·47 | 0·99 (0·93 to 1·05) | 26,930 (3·41) | 10·73 | 0·84 (0·81 to 0·88) | 16,785 (1·37) | 8·58 | 0·90 (0·85 to 0·95) |
| South Asian | 25,240 (6·08) | 12·08 | 0·74 (0·72 to 0·77) | 71,565 (9·06) | 7·41 | 0·57 (0·55 to 0·59) | 58,635 (4·79) | 5·98 | 0·63 (0·61 to 0·65) |
| Mixed | 6,450 (1·55) | 14·42 | 0·94 (0·87 to 1·00) | 9,520 (1·21) | 10·71 | 0·87 (0·81 to 0·93) | 6,220 (0·51) | 8·52 | 0·94 (0·86 to 1·03) |
| Chinese/Other | 10,555 (2·54) | 12·41 | 0·79 (0·74 to 0·83) | 31,245 (3·96) | 7·91 | 0·63 (0·60 to 0·65) | 24,595 (2·01) | 6·32 | 0·69 (0·65 to 0·72) |
| **Patient IMD** | | | | | | | | | |
| 1 | 107,900 (26·00) | 16·46 | 1 | 200,040 (25·33) | 12·35 | 1 | 233,525 (19·10) | 9·60 | 1 |
| 2 | 90,885 (21·90) | 15·22 | 0·90 (0·88 to 0·92) | 171,415 (21·71) | 11·74 | 0·93 (0·91 to 0·95) | 241,715 (19·77) | 8·89 | 0·92 (0·90 to 0·94) |
| 3 | 83,715 (20·17) | 14·11 | 0·82 (0·80 to 0·84) | 161,950 (20·51) | 11·11 | 0·85 (0·83 to 0·86) | 272,565 (22·29) | 8·35 | 0·85 (0·83 to 0·87) |
| 4 | 72,925 (17·57) | 13·49 | 0·77 (0·75 to 0·79) | 138,800 (17·58) | 10·43 | 0·77 (0·75 to 0·79) | 252,180 (20·62) | 7·86 | 0·79 (0·77 to 0·81) |
| 5 | 59,650 (14·37) | 12·46 | 0·70 (0·68 to 0·72) | 117,490 (14·88) | 9·54 | 0·69 (0·67 to 0·71) | 222,965 (18·23) | 7·44 | 0·74 (0·73 to 0·76) |
| **Long Term Condition** | | | | | | | | | |
| Hypertension | | | | | | | | | |
| Absent | 394,430 (95·03) | 14·58 | 1 | 504,485 (63·88) | 11·17 | 1 | 400,210 (32·73) | 8·36 | 1 |
| Present | 20,640 (4·97) | 15·26 | 1·07 (1·03 to 1·12) | 285,215 (36·12) | 11·28 | 1·06 (1·04 to 1·08) | 822,740 (67·27) | 8·46 | 1·03 (1·01 to 1·04) |
| Type 1 Diabetes | | | | | | | | | |
| Absent | 403,630 (97·24) | 14·67 | 1 | 769,935 (97·50) | 11·25 | 1 | 1,208,365 (98·81) | 8·45 | 1 |
| Present | 11,445 (2·76) | 12·76 | 0·88 (0·83 to 0·93) | 19,765 (2·50) | 9·41 | 0·84 (0·80 to 0·88) | 14,585 (1·19) | 7·10 | 0·84 (0·79 to 0·89) |
| Type 2 Diabetes | | | | | | | | | |
| Absent | 397,320 (95·72) | 14·66 | 1 | 584,335 (74·00) | 11·41 | 1 | 766,865 (62·71) | 8·31 | 1 |
| Present | 17,755 (4·28) | 13·69 | 0·95 (0·91 to 1·00) | 205,360 (26·00) | 10·62 | 1·02 (1·00 to 1·03) | 456,085 (37·29) | 8·64 | 1·11 (1·09 to 1·12) |
| Cardiovascular Disease | | | | | | | | | |
| Absent | 409,100 (98·56) | 14·61 | 1 | 725,700 (91·90) | 11·16 | 1 | 925,885 (75·71) | 8·44 | 1 |
| Present | 5,970 (1·44) | 15·08 | 1·05 (0·98 to 1·13) | 63,995 (8·10) | 11·75 | 1·12 (1·09 to 1·15) | 297,065 (24·29) | 8·38 | 1·06 (1·04 to 1·07) |
| Learning Disability | | | | | | | | | |
| :Absent | 395,630 (95·32) | 14·62 | 1 | 769,390 (97·43) | 11·15 | 1 | 1,213,315 (99·21) | 8·39 | 1 |
| Present | 19,445 (4·68) | 14·53 | 1·02 (0·98 to 1·07) | 20,310 (2·57) | 13·22 | 1·19 (1·14 to 1·24) | 9,635 (0·79) | 13·91 | 1·71 (1·61 to 1·81) |
| Depression | | | | | | | | | |
| :Absent | 292,005 (70·35) | 13·36 | 1 | 511,055 (64·72) | 9·93 | 1 | 905,980 (74·08) | 7·79 | 1 |
| Present | 123,070 (29·65) | 17·61 | 1·34 (1·32 to 1·37) | 278,645 (35·28) | 13·55 | 1·31 (1·29 to 1·33) | 316,970 (25·92) | 10·25 | 1·24 (1·23 to 1·26) |
| Dementia | | | | | | | | | |
| :Absent | 414,955 (99·97) | 14·62 | 1 | 787,345 (99·70) | 11·20 | 1 | 1,201,495 (98·25) | 8·36 | 1 |
| Present | 120 (0·03) | 16·67 | 1·11 (0·66 to 1·77) | 2,350 (0·30) | 12·77 | 1·15 (1·02 to 1·30) | 21,460 (1·75) | 12·12 | 1·51 (1·44 to 1·57) |
| Serious Mental Illness | | | | | | | | | |
| Absent | 400,270 (96·43) | 14·47 | 1 | 749,590 (94·92) | 11·01 | 1 | 1,193,430 (97·59) | 8·32 | 1 |
| :Present | 14,805 (3·57) | 18·47 | 1·38 (1·32 to 1·44) | 40,105 (5·08) | 14·87 | 1·41 (1·37 to 1·45) | 29,520 (2·41) | 12·79 | 1·54 (1·49 to 1·60) |
| Asthma | | | | | | | | | |
| Absent | 296,605 (71·46) | 14·21 | 1 | 587,935 (74·45) | 10·89 | 1 | 967,685 (79·13) | 8·20 | 1 |
| Present | 118,465 (28·54) | 15·63 | 1·11 (1·09 to 1·13) | 201,760 (25·55) | 12·13 | 1·08 (1·07 to 1·10) | 255,270 (20·87) | 9·29 | 1·09 (1·07 to 1·11) |
| COPD | | | | | | | | | |
| Absent | 413,490 (99·62) | 14·62 | 1 | 753,610 (95·43) | 11·10 | 1 | 1,063,920 (87·00) | 8·27 | 1 |
| Present | 1,585 (0·38) | 14·51 | 0·96 (0·83 to 1·10) | 36,085 (4·57) | 13·54 | 1·16 (1·13 to 1·20) | 159,030 (13·00) | 9·52 | 1·11 (1·09 to 1·13) |

| | | | | | | | | | |
|---|---|---|---|---|---|---|---|---|---|
| Stroke and TIA | | | | | | | | | |
| Absent | 413,265 (99·56) | 14·61 | 1 | 767,285 (97·16) | 11·17 | 1 | 1,115,200 (91·19) | 8·38 | 1 |
| Present | 1,810 (0·44) | 16·02 | 1·12 (0·99 to 1·27) | 22,415 (2·84) | 12·45 | 1·13 (1·08 to 1·17) | 107,755 (8·81) | 8·96 | 1·09 (1·07 to 1·12) |

δ-change: This refers to the change (δ) in rate of weight gain after the onset of the pandemic (δ-change = δ-pandemic - δ-prepandemic)
AR: refers to the absolute risk of experiencing a extreme acceleration in rate of weight gain
aOR: Adjusted for sex, individual Index of Multiple Deprivation and ethnicity
Serious Mental Illness includes psychosis and bipolar disorder
Chronic Obstructive Pulmonary Disease (COPD)

#### Supplementary Table 4: Ethnicity stratified associations with risk of extreme acceleration in rate of weight gain

**Supplementary Table 4: Ethnicity stratified analysis of associations between sociodemographic and clinical characteristics and risk of extreme acceleration in rate of weight gain (δ-change >= 1·84 kg/m²/year) during the pandemic amongst Black and South Asian (Indian, Pakistani, Bangladeshi) individuals**

| | **South Asian** | | | **Black** | | |
|---|---|---|---|---|---|---|
| | **N (%)** | **AR (%)** | **aOR (95% C)** | **N (%)** | **AR (%)** | **aOR (95% C)** |
| **Sex** | | | | | | |
| F | 92,185 (55·78) | 8·77 | 1 | 33,525 (60·29) | 12·11 | 1 |
| M | 73,080 (44·22) | 6·10 | 0·73 (0·71 to 0·76) | 22,080 (39·71) | 8·11 | 0·69 (0·65 to 0·73) |
| **Age Group (years)** | | | | | | |
| 18-29 | 6,055 (3·66) | 13·71 | 1 | 2,040 (3·67) | 16·91 | 1 |
| 30-39 | 19,185 (11·61) | 11·57 | 0·83 (0·76 to 0·90) | 5,495 (9·88) | 14·92 | 0·86 (0·75 to 0·99) |
| 40-49 | 35,395 (21·42) | 8·12 | 0·59 (0·54 to 0·64) | 10,625 (19·11) | 11·86 | 0·69 (0·61 to 0·79) |
| 50-59 | 36,170 (21·89) | 6·70 | 0·49 (0·45 to 0·53) | 16,305 (29·33) | 10·00 | 0·59 (0·52 to 0·67) |
| 60-69 | 37,785 (22·86) | 6·01 | 0·44 (0·40 to 0·47) | 11,060 (19·89) | 8·68 | 0·51 (0·45 to 0·58) |
| 70-79 | 20,850 (12·62) | 5·90 | 0·42 (0·39 to 0·47) | 5,725 (10·30) | 8·30 | 0·48 (0·41 to 0·55) |
| 80 | 9,830 (5·95) | 6·97 | 0·51 (0·46 to 0·57) | 4,355 (7·83) | 8·15 | 0·48 (0·41 to 0·56) |
| **Patient IMD** | | | | | | |
| 1 | 62,560 (37·85) | 8·15 | 1 | 23,530 (42·32) | 10·96 | 1 |
| 2 | 46,570 (28·18) | 7·42 | 0·95 (0·91 to 0·99) | 14,760 (26·55) | 10·26 | 0·94 (0·88 to 1·00) |
| 3 | 29,980 (18·14) | 7·25 | 0·94 (0·89 to 0·99) | 9,260 (16·65) | 10·26 | 0·96 (0·89 to 1·04) |
| 4 | 15,615 (9·45) | 7·04 | 0·91 (0·85 to 0·98) | 5,140 (9·25) | 9·34 | 0·87 (0·78 to 0·96) |
| 5 | 10,545 (6·38) | 6·78 | 0·89 (0·82 to 0·96) | 2,910 (5·23) | 11·00 | 1·04 (0·92 to 1·18) |
| **Long Term Condition** | | | | | | |
| Hypertension | | | | | | |
| Absent | 89,415 (54·10) | 8·41 | 1 | 26,685 (47·99) | 11·67 | 1 |
| Present | 75,855 (45·90) | 6·62 | 1·00 (0·96 to 1·05) | 28,915 (52·01) | 9·46 | 1·00 (0·94 to 1·07) |
| Type 1 Diabetes | | | | | | |
| Absent | 163,960 (99·21) | 7·58 | 1 | 54,775 (98·51) | 10·53 | 1 |
| Present | 1,310 (0·79) | 8·02 | 0·95 (0·77 to 1·15) | 825 (1·49) | 9·70 | 0·90 (0·70 to 1·13) |
| Type 2 Diabetes | | | | | | |
| Absent | 81,205 (49·14) | 8·95 | 1 | 33,200 (59·71) | 11·67 | 1 |
| Present | 84,060 (50·86) | 6·27 | 0·84 (0·80 to 0·87) | 22,400 (40·29) | 8·82 | 0·88 (0·83 to 0·94) |
| Cardiovascular Disease | | | | | | |
| Absent | 139,690 (84·52) | 7·72 | 1 | 50,115 (90·13) | 10·57 | 1 |
| Present | 25,575 (15·47) | 6·88 | 1·16 (1·10 to 1·23) | 5,485 (9·87) | 10·03 | 1·19 (1·08 to 1·31) |
| Learning Disability | | | | | | |
| Absent | 162,845 (98·53) | 7·53 | 1 | 54,840 (98·63) | 10·49 | 1 |
| Present | 2,425 (1·47) | 11·13 | 1·16 (1·01 to 1·32) | 760 (1·37) | 12·50 | 1·06 (0·84 to 1·31) |
| Depression | | | | | | |
| Absent | 133,545 (80·81) | 7·21 | 1 | 44,725 (80·44) | 10·03 | 1 |
| Present | 31,725 (19·20) | 9·16 | 1·26 (1·21 to 1·32) | 10,875 (19·56) | 12·51 | 1·22 (1·14 to 1·30) |
| Dementia | | | | | | |
| Absent | 162,655 (98·42) | 7·58 | 1 | 54,625 (98·24) | 10·50 | 1 |
| Present | 2,610 (1·58) | 8·05 | 1·27 (1·10 to 1·47) | 975 (1·76) | 11·79 | 1·48 (1·20 to 1·81) |
| Serious Mental Illness | | | | | | |
| Absent | 158,655 (96·00) | 7·39 | 1 | 51,835 (93·23) | 10·15 | 1 |
| Present | 6,610 (4·00) | 12·25 | 1·68 (1·55 to 1·81) | 3,765 (6·77) | 15·67 | 1·63 (1·48 to 1·79) |
| Asthma | | | | | | |
| Absent | 131,465 (79·55) | 7·45 | 1 | 45,075 (81·07) | 10·32 | 1 |
| Present | 33,805 (20·45) | 8·13 | 1·09 (1·04 to 1·14) | 10,525 (18·93) | 11·40 | 1·08 (1·01 to 1·16) |
| COPD | | | | | | |
| Absent | 160,075 (96·86) | 7·60 | 1 | 53,955 (97·04) | 10·53 | 1 |
| Present | 5,190 (3·14) | 7·23 | 1·19 (1·07 to 1·33) | 1,645 (2·96) | 10·33 | 1·22 (1·04 to 1·44) |
| Stroke and TIA | | | | | | |
| Absent | 157,710 (95·42) | 7·57 | 1 | 53,085 | 10·6 | 1 |
| Present | 7,560 (4·57) | 7·87 | 1·28 (1·18 to 1·40) | 2520 | 9·52 | 1·09 (0·94 to 1·25) |

δ-change: This refers to the change (δ) in rate of weight gain after the onset of the pandemic (δ-change = δ-pandemic - δ-prepandemic)
AR: refers to the absolute risk of experiencing extreme acceleration in rate of weight gain
aOR: Adjusted for age, sex and individual Index of Multiple Deprivation
Serious Mental Illness includes psychosis and bipolar disorder
Chronic Obstructive Pulmonary Disease (COPD)

#### Supplementary Table 5: Sex stratified associations with risk of extreme acceleration in rate of weight gain

**Supplementary Table 5: Sex stratified analysis of associations between sociodemographic and clinical characteristics and risk of extreme acceleration in rate of weight gain (δ-change >= 1·84 kg/m²/year) during the pandemic**

| | Female | | | Male | | |
|---|---|---|---|---|---|---|
| | N (%) | AR (%) | aOR(95% CI) | N (%) | AR (%) | aOR(95% CI) |
| Sex | | | | | | |
| **Age Group (years)** | | | | | | |
| 18-29 | 145,605 (9·03) | 14·86 | 1 | 16,050 (1·39) | 14·89 | 1 |
| 30-39 | 215,520 (13·36) | 14·71 | 1·00 (0·98 to 1·02) | 37,895 (3·28) | 13·05 | 0·87 (0·83 to 0·92) |
| 40-49 | 214,595 (13·30) | 12·51 | 0·85 (0·83 to 0·86) | 95,450 (8·26) | 10·36 | 0·69 (0·66 to 0·72) |
| 50-59 | 263,900 (16·36) | 12·10 | 0·80 (0·79 to 0·82) | 215,755 (18·67) | 9·20 | 0·58 (0·56 to 0·61) |
| 60-69 | 273,820 (16·98) | 11·00 | 0·72 (0·71 to 0·74) | 299,215 (25·89) | 7·71 | 0·48 (0·46 to 0·50) |
| 70-79 | 315,985 (19·59) | 9·18 | 0·59 (0·58 to 0·60) | 333,935 (28·89) | 6·25 | 0·38 (0·36 to 0·40) |
| 80 | 183,430 (11·37) | 8·50 | 0·54 (0·53 to 0·56) | 157,545 (13·63) | 5·72 | 0·35 (0·33 to 0·37) |
| **Ethnicity** | | | | | | |
| White | 1,432,010 (88·79) | 11·82 | 1 | 22,080 (1·91) | 8·11 | 1 |
| Black | 33,525 (2·08) | 12·11 | 0·92 (0·89 to 0·95) | 30,345 (2·62) | 6·69 | 0·83 (0·79 to 0·87) |
| South Asian | 92,185 (5·72) | 8·77 | 0·64 (0·63 to 0·66) | 8,050 (0·70) | 8·82 | 0·60 (0·58 to 0·62) |
| Mixed | 15,240 (0·95) | 12·24 | 0·91 (0·86 to 0·95) | 73,080 (6·32) | 6·10 | 0·90 (0·83 to 0·97) |
| Chinese/Other | 39,890 (2·47) | 8·89 | 0·67 (0·65 to 0·70) | 1,022,295 (88·45) | 7·93 | 0·69 (0·65 to 0·72) |
| IMD | | | | | | |
| 1 | 355,210 (22·02) | 13·38 | 1 | 237,595 (20·56) | 9·11 | 1 |
| 2 | 332,785 (20·63) | 12·26 | 0·91 (0·90 to 0·93) | 232,440 (20·11) | 8·32 | 0·93 (0·91 to 0·95) |
| 3 | 344,265 (21·34) | 11·30 | 0·84 (0·82 to 0·85) | 252,485 (21·84) | 7·62 | 0·86 (0·84 to 0·88) |
| 4 | 310,370 (19·24) | 10·65 | 0·78 (0·77 to 0·79) | 230,750 (19·96) | 7·09 | 0·80 (0·78 to 0·82) |
| 5 | 270,215 (16·75) | 9·83 | 0·71 (0·70 to 0·73) | 202,585 (17·53) | 6·62 | 0·75 (0·73 to 0·77) |
| **Long Term Condition** | | | | | | |
| Hypertension | | | | | | |
| Absent | 925,485 (57·38) | 12·40 | 1 | 443,910 (38·41) | 8·43 | 1 |
| Present | 687,365 (42·62) | 10·48 | 1·08 (1·07 to 1·10) | 711,935 (61·59) | 7·39 | 1·03 (1·02 to 1·05) |
| Type 1 Diabetes | | | | | | |
| Absent | 1,591,355 (98·67) | 11·58 | 1 | 1,129,435 (97·72) | 7·79 | 1 |
| Present | 21,495 (1·33) | 11·61 | 0·94 (0·91 to 0·98) | 26,410 (2·28) | 7·57 | 0·74 (0·71 to 0·78) |
| Type 2 Diabetes | | | | | | |
| Absent | 1,257,425 (77·96) | 11·79 | 1 | 705,950 (61·08) | 7·89 | 1 |
| Present | 355,425 (22·04) | 10·87 | 1·08 (1·07 to 1·10) | 449,895 (38·92) | 7·63 | 1·06 (1·04 to 1·07) |
| Cardiovascular Disease | | | | | | |
| Absent | 1,424,260 (88·31) | 11·69 | 1 | 846,130 (73·20) | 7·94 | 1 |
| Present | 188,590 (11·69) | 10·78 | 1·13 (1·11 to 1·14) | 309,715 (26·80) | 7·37 | 1·09 (1·07 to 1·11) |
| Learning Disability | | | | | | |
| Absent | 1,590,870 (98·64) | 11·53 | 1 | 1,127,955 (97·59) | 7·67 | 1 |
| Present | 21,980 (1·36) | 15·65 | 1·22 (1·18 to 1·27) | 27,890 (2·41) | 12·51 | 1·15 (1·10 to 1·19) |
| Depression | | | | | | |
| Absent | 1,081,410 (67·05) | 10·54 | 1 | 909,935 (78·72) | 7·23 | 1 |
| Present | 531,440 (32·95) | 13·70 | 1·28 (1·27 to 1·30) | 245,910 (21·28) | 9·87 | 1·26 (1·24 to 1·28) |
| Dementia | | | | | | |
| Absent | 1,586,340 (98·36) | 11·53 | 1 | 1,133,945 (98·11) | 7·76 | 1 |
| Present | 26,510 (1·64) | 14·65 | 1·76 (1·70 to 1·83) | 21,900 (1·90) | 9·38 | 1·55 (1·48 to 1·62) |
| SMI | | | | | | |
| Absent | 1,565,370 (97·06) | 11·45 | 1 | 1,114,950 (96·46) | 7·60 | 1 |
| Present | 47,480 (2·94) | 16·10 | 1·44 (1·40 to 1·48) | 40,895 (3·54) | 13·00 | 1·45 (1·41 to 1·49) |
| Asthma | | | | | | |
| Absent | 1,201,175 (74·48) | 11·18 | 1 | 929,035 (80·38) | 7·63 | 1 |
| Present | 411,675 (25·52) | 12·75 | 1·11 (1·10 to 1·13) | 226,810 (19·62) | 8·46 | 1·03 (1·01 to 1·05) |
| COPD | | | | | | |
| Absent | 1,494,310 (92·65) | 11·58 | 1 | 1,028,875 (89·02) | 7·75 | 1 |
| Present | 118,540 (7·35) | 11·68 | 1·13 (1·11 to 1·15) | 126,970 (10·98) | 8·13 | 1·16 (1·13 to 1·18) |
| Stroke and TIA | | | | | | |
| Absent | 1,526,200 (94·63) | 11·62 | 1 | 1,051,995 (91·02) | 7·82 | 1 |
| Present | 86,650 (5·37) | 10·88 | 1·13 (1·10 to 1·16) | 103,850 (8·98) | 7·53 | 1·12 (1·09 to 1·14) |

δ-change: This refers to the change (δ) in rate of weight gain after the onset of the pandemic (δ-change = δ-pandemic - δ-prepandemic)
AR: refers to the absolute risk of experiencing extreme acceleration in rate of weight gain
aOR: Adjusted for age, ethnicity and individual Index of Multiple Deprivation
Serious Mental Illness includes psychosis and bipolar disorder
Chronic Obstructive Pulmonary Disease (COPD)

#### Supplementary Table 6: Index of Multiple Deprivation (IMD) stratified associations with risk of extreme acceleration in rate of weight gain

**Supplementary Table 6: Analysis of associations between sociodemographic and clinical characteristics and risk of extreme acceleration in rate of weight gain (δ-change >= 1·84 kg/m$^2$/year) during the pandemic stratified by individual level Index of Multiple Deprivation (IMD)**

| | IMD 1 (most deprived) | | | IMD 5 (least deprived) | | |
|---|---|---|---|---|---|---|
| | N (%) | AR (%) | aOR (95% CI) | N (%) | AR (%) | aOR (95% CI) |
| **Sex** | | | | | | |
| F | 355,210 (59·92) | 13·38 | 1 | 270,215 (57·15) | 9·83 | 1 |
| M | 237,595 (40·08) | 9·11 | 0·73 (0·71 to 0·74) | 202,585 (42·85) | 6·62 | 0·72 (0·70 to 0·74) |
| **Age Group (years)** | | | | | | |
| 18-29 | 39,950 (6·74) | 17·46 | 1 | 24,620 (5·21) | 12·06 | 1 |
| 30-39 | 67,950 (11·46) | 15·87 | 0·94 (0·91 to 0·97) | 5,030 (7·41) | 12·75 | 1·08 (1·03 to 1·14) |
| 40-49 | 81,210 (13·70) | 13·04 | 0·81 (0·78 to 0·84) | 45,505 (9·62) | 9·97 | 0·86 (0·82 to 0·90) |
| 50-59 | 118,835 (20·05) | 11·88 | 0·73 (0·70 to 0·75) | 71,985 (15·22) | 9·27 | 0·84 (0·80 to 0·88) |
| 60-69 | 124,975 (21·08) | 10·27 | 0·62 (0·60 to 0·64) | 94,455 (19·98) | 8·21 | 0·76 (0·72 to 0·79) |
| 70-79 | 108,550 (18·31) | 8·83 | 0·51 (0·49 to 0·52) | 128,515 (27·18) | 6·88 | 0·62 (0·59 to 0·65) |
| 80 | 51,340 (8·66) | 8·36 | 0·47 (0·45 to 0·49) | 72,695 (15·38) | 6·55 | 0·58 (0·55 to 0·61) |
| **Ethnicity** | | | | | | |
| White | 480,580 (81·07) | 12·24 | 1 | 449,885 (95·15) | 8·51 | 1 |
| Black | 23,530 (3·97) | 10·96 | 0·84 (0·81 to 0·88) | 2,910 (0·61) | 11·00 | 1·24 (1·10 to 1·39) |
| South Asian | 62,560 (10·55) | 8·15 | 0·60 (0·58 to 0·62) | 10,545 (2·23) | 6·78 | 0·75 (0·69 to 0·81) |
| Mixed | 7,175 (1·21) | 12·54 | 0·90 (0·84 to 0·96) | 2,505 (0·53) | 8·58 | 0·87 (0·75 to 1·00) |
| Chinese/Other | 18,960 (3·20) | 9·20 | 0·69 (0·66 to 0·73) | 6,955 (1·47) | 6·76 | 0·73 (0·67 to 0·81) |
| **Long Term Condition** | | | | | | |
| Hypertension | | | | | | |
| Absent | 310,530 (52·38) | 12·94 | 1 | 226,165 (47·84) | 9·22 | 1 |
| Present | 282,275 (47·62) | 10·27 | 1·04 (1·02 to 1·06) | 246,635 (52·16) | 7·76 | 1·12 (1·09 to 1·15) |
| Type 1 Diabetes | | | | | | |
| Absent | 584,060 (98·52) | 11·66 | 1 | 463,390 (98·01) | 8·47 | 1 |
| Present | 8,745 (1·47) | 11·84 | 0·94 (0·88 to 1·00) | 9,410 (1·99) | 8·02 | 0·90 (0·84 to 0·97) |
| Type 2 Diabetes | | | | | | |
| Absent | 402,955 (67·97) | 12·52 | 1 | 350,905 (74·22) | 8·57 | 1 |
| Present | 189,850 (32·03) | 9·86 | 0·97 (0·95 to 0·99) | 121,895 (25·78) | 8·14 | 1·17 (1·15 to 1·20) |
| Cardiovascular Disease | | | | | | |
| Absent | 487,765 (82·28) | 12·02 | 1 | 386,645 (81·78) | 8·70 | 1 |
| Present | 105,040 (17·72) | 10·04 | 1·08 (1·05 to 1·10) | 86,155 (18·22) | 7·38 | 1·09 (1·06 to 1·13) |
| Learning Disability | | | | | | |
| Absent | 577,975 (97·50) | 11·60 | 1 | 467,665 (98·91) | 8·42 | 1 |
| Present | 14,830 (2·50) | 14·36 | 1·13 (1·08 to 1·19) | 5,135 (1·09) | 11·88 | 1·28 (1·17 to 1·39) |
| Depression | | | | | | |
| Absent | 393,500 (66·38) | 10·55 | 1 | 361,765 (76·52) | 7·71 | 1 |
| Present | 199,305 (33·62) | 13·88 | 1·23 (1·21 to 1·25) | 111,035 (23·48) | 10·89 | 1·36 (1·33 to 1·39) |
| Dementia | | | | | | |
| Absent | 582,520 (98·27) | 11·64 | 1 | 464,200 (98·18) | 8·41 | 1 |
| Present | 10,285 (1·74) | 13·37 | 1·59 (1·50 to 1·69) | 8,600 (1·82) | 11·22 | 1·75 (1·63 to 1·88) |
| SMI | | | | | | |
| Absent | 565,585 (95·41) | 11·48 | 1 | 463,300 (97·99) | 8·36 | 1 |
| Present | 27,220 (4·59) | 15·48 | 1·40 (1·35 to 1·45) | 9,500 (2·01) | 13·26 | 1·60 (1·50 to 1·69) |
| Asthma | | | | | | |
| Absent | 442,175 (74·59) | 11·18 | 1 | 370,555 (78·37) | 8·23 | 1 |
| Present | 150,630 (25·41) | 13·11 | 1·12 (1·10 to 1·14) | 102,245 (21·63) | 9·29 | 1·07 (1·05 to 1·10) |
| COPD | | | | | | |
| Absent | 520,565 (87·81) | 11·78 | 1 | 443,150 (93·73) | 8·45 | 1 |
| Present | 72,240 (12·19) | 10·86 | 1·09 (1·06 to 1·12) | 29,650 (6·27) | 8·60 | 1·24 (1·18 to 1·29) |
| Stroke and TIA | | | | | | |
| Absent | 553,750 (93·41) | 11·74 | 1 | 439,290 (92·91) | 8·50 | 1 |
| Present | 39,055 (6·59) | 10·66 | 1·13 (1·09 to 1·16) | 33,510 (7·09) | 7·94 | 1·16 (1·11 to 1·21) |

δ-change: This refers to the change (δ) in rate of weight gain after the onset of the pandemic (δ-change = δ-pandemic - δ-prepandemic)
AR: refers to the absolute risk of experiencing extreme acceleration in rate of weight gain
aOR: Adjusted for age, ethnicity and individual Index of Multiple Deprivation
Serious Mental Illness includes Psychosis and Bipolar Disorder
Chronic Obstructive Pulmonary Disease (COPD)
